# Specific immune-regulatory transcriptional signatures reveal sex and age differences in SARS-CoV-2 infected patients

**DOI:** 10.1101/2020.11.12.20230417

**Authors:** Paula Paccielli Freire, Alexandre H. C. Marques, Gabriela Crispim Baiocchi, Lena F. Schimke, Dennyson Leandro M. Fonseca, Ranieri Coelho Salgado, Igor Salerno Filgueiras, Sarah Maria da Silva Napoleao, Desirée Rodrigues Plaça, Thiago Dominguez Crespo Hirata, Nadia El Khawanky, Lasse Melvaer Giil, Gustavo Cabral de Miranda, Robson Francisco Carvalho, Luis Carlos de Souza Ferreira, Antonio Condino-Neto, Helder Takashi Imoto Nakaya, Igor Jurisica, Hans D. Ochs, Niels Olsen Saraiva Camara, Vera Lúcia Garcia Calich, Otavio Cabral-Marques

**Affiliations:** Department of Immunology, Institute of Biomedical Sciences, University of São Paulo, São Paulo, SP, Brazil; Department of Clinical and Toxicological Analyses, School of Pharmaceutical Sciences, University of São Paulo, São Paulo, SP, Brazil; Department of Hematology and Oncology, Faculty of Medicine, The University of Freiburg, Freiburg, Germany; Department of Internal Medicine, Haraldsplass Deaconess Hospital, Bergen, Norway; Department of Structural and Functional Biology, Institute of Biosciences, São Paulo State University (UNESP), Botucatu, SP, Brazil; Laboratório de Desenvolvimento de Vacinas, Instituto de Ciências Biomédicas, Departamento de Microbiologia, Universidade de São Paulo, São Paulo, SP, Brazil; Krembil Research Institute, UHN; and Departments of Medical Biophysics and Computer Science, University of Toronto, Toronto, Canada; Department of Pediatrics, University of Washington School of Medicine, and Seattle Children’s Research Institute, Seattle, WA; Network of Immunity in Infection, Malignancy, and Autoimmunity (NIIMA), Universal Scientific Education and Research Network (USERN), Sao Paulo, Brazil

## Abstract

The coronavirus disease 2019 (COVID-19) fatality rate varies in different patient groups. However, the underlying mechanisms that explain this variation are poorly understood. Here, we reanalyzed and integrated public RNAseq datasets of nasopharyngeal swabs and peripheral blood leukocytes from patients with SARS-CoV-2, comparing transcription patterns according to sex, age, and viral load. We found that female and young patients infected by SARS-CoV-2 exhibited a similar transcriptomic pattern with a larger number of total (up- and downregulated) differentially expressed genes (DEGs) compared to males and elderly patients. The transcriptional analysis showed a sex-specific profile with a higher transcriptional modulation of immune response-associated genes in female and young subjects against SARS-CoV-2. The functional clustering was characterized by a highly correlated interferome network of cytokine/chemokine- and neutrophil-associated genes that were enriched both in nasopharyngeal cells and peripheral blood of COVID-19 patients. Females exhibited reduced transcriptional levels of key pro-inflammatory/neutrophil-related genes such as CXCL8 receptors (*CXCR1/CXCR2*), *IL-1β, S100A9, ITGAM*, and *DBNL* compared to males, which correlate with a protective gene expression profile against inflammatory damage. Our data indicate specific immune-regulatory pathways associated with sex and age of patients infected with SARS-CoV-2. These results point out therapeutic targets to reduce morbidity and mortality of COVID-19.

## INTRODUCTION

More than ten months after the outbreak of the novel Coronavirus disease 2019 (COVID-19) in Wuhan, China^1–3^, approximately 43 million confirmed cases of COVID-19 and more than 1,1 million deaths have been reported worldwide^4^. The clinical spectrum of COVID-19 ranges from asymptomatic to severe pulmonary disease, leading to acute respiratory distress syndrome (ARDS) ^5,6^. Enhanced expression of the angiotensin-converting enzyme 2 (ACE2), the SARS-CoV-2 entry receptor^7^, and dysregulation of the immune response likely contribute to more severe disease in older patients with comorbidities. Even as there is an increased understanding of what increases the risk of severe disease, the increased male/female mortality ratio remains poorly understood, especially considering the lack of gender differences in disease incidence^6,8^. A recent study reported no gender differences in the levels of anti-S1-IgM and -IgG antibody titers nor the number of naïve- or memory B cells. Compared to males, peripheral blood mononuclear cells (PBMCs) from females exhibited higher levels of terminally differentiated T cells expressing activation molecules (CD38 and HLA-DR-positive) and negative regulators (PD-1 and TIM-3) ^9^. Moreover, male patients displayed higher plasma levels of pro-inflammatory innate immune cytokine/chemokines (*CXCL8* and *IL-18*)^10^. These findings indicate a better capacity for immune modulation in females compared to males.

The immune response to SARS-CoV-2 is characterized by hyperactivated T cells (both CD4^+^ and CD8^+^) ^6,11^ and macrophages ^12^. These hyperactivated immune cells likely contribute to the massive serum levels of pro-inflammatory cytokines, also referred to as “cytokine storm” ^13^. In parallel, tissue damage has been associated with hyperactivation of neutrophil-induced oxidative stress and high neutrophil counts^14^, degranulation, and release of extracellular traps (NETs). These processes are associated with increased levels of acute-phase reactants (e.g., C-reactive protein), microvascular damage, arterial thrombosis, and red blood cell dysfunction ^14,15^. Moreover, an association between the occurrence of neutrophilia and increased production of granulocytic myeloid-derived suppressor cells (MDSCs) has also been reported. However, it is not clear whether this is harmful or adaptive ^16–18^. Whether the function of the MDSC is different between men and women or young and elderly with COVID-19 remains an open issue.

Several omic studies have been conducted in patients with COVID-19 to help decipher the molecular mechanisms underlying the disease. Lieberman et al. 2020, ^19^ and Mick et al., 2020 ^20^, recently performed RNASeq experiments investigating global transcriptional profiles of nasopharyngeal swabs from 668 individuals with SARS-CoV-2 (SC2) and 157 individuals negative of SARS-CoV-2 (Neg. SC2). Although these studies have highlighted differences in immune responses that underlie disparities in male and elderly outcomes, a consensus profile reporting the cytokines and genes associated with neutrophil-mediated immunity is still missing. The specific set of immune system genes in females underlying the protective mechanisms is another question that must be better understood and discussed. Here, we characterize a previously unnoticed interconnected transcriptome network between differentially expressed genes (DEGs) associated with cytokine-mediated signaling pathway (CMSP) and neutrophil mediated immunity (NMI) genes accordingly to sex and age. We found specific transcriptome signatures in RNASeq from nasopharyngeal swabs and peripheral blood leukocytes of human samples (**Supplementary Table 1**). Finally, we sought to identify immune transcriptomic profiles according to sex and age, as well as the regulatory networks governing the immune response to SARS-CoV-2.

## RESULTS

### Association between cytokine/chemokine- and neutrophil-related genes in nasopharyngeal swabs

We first performed a transcriptomic reanalysis of nasopharyngeal swabs from patients described by Lieberman et al., 2020 (GSE152075)^19^ to characterize the expression landscape of immune system genes. We divided the samples according to sex, age (elderly> 60y and young <60 y), and viral load (413 patients infected with SARS-CoV-2 and 54 negative controls) (**Fig. 1a)**. Groups of patients with high and low viral loads, and younger and older age, were gender-matched to avoid confounding effects (**Supplementary Tables S1 and S2**). Female and young patients infected by SARS-CoV-2 exhibited a higher number of total (up- and down-regulated) differentially expressed genes (DEGs) when compared to male and elderly patients (**Fig. 1b, Supplementary Table S3**). Genes significantly deregulated in each group of patients were selected to perform enrichment analysis using the Gene Ontology terms through the Enrichr tool ^21,22^. Overall, we observed cytokine-mediated signaling pathway (CMSP) and neutrophil-mediated immunity (NMI) categories were enriched by up- and down-regulated genes, respectively, among the compiled top-10 biological process categories of each group (**Supplementary Tables S4** and **S5**). Also, we used CEMiTool^23^ to gain insights into the systemic function of nasopharyngeal swab genes by performing modular co-expression enrichment and network analyses. Among the gene-modules identified, module 1 (M1) indicated a co-expression association between genes associated with neutrophil degranulation signaling by interleukins and GPCR ligand binding (chemokines and their receptors) (**Fig. 1c**). When we compared the total number of genes associated with CMSP and NMI categories, female and young patients present higher numbers of up- and down-regulated DEGs associated with these categories compared to male and elderly patients (**Fig. 1d and Supplementary Tables S6 and S7**). Females displayed an enhanced quantity of DEGs compared to male, elderly, and patients with low viral load (described in **Supplementary Tables S6-S8**). These observations indicate a higher transcriptional modulation of the immune response in female and young patients compared to male and elderly patients.

**Fig. 1.**
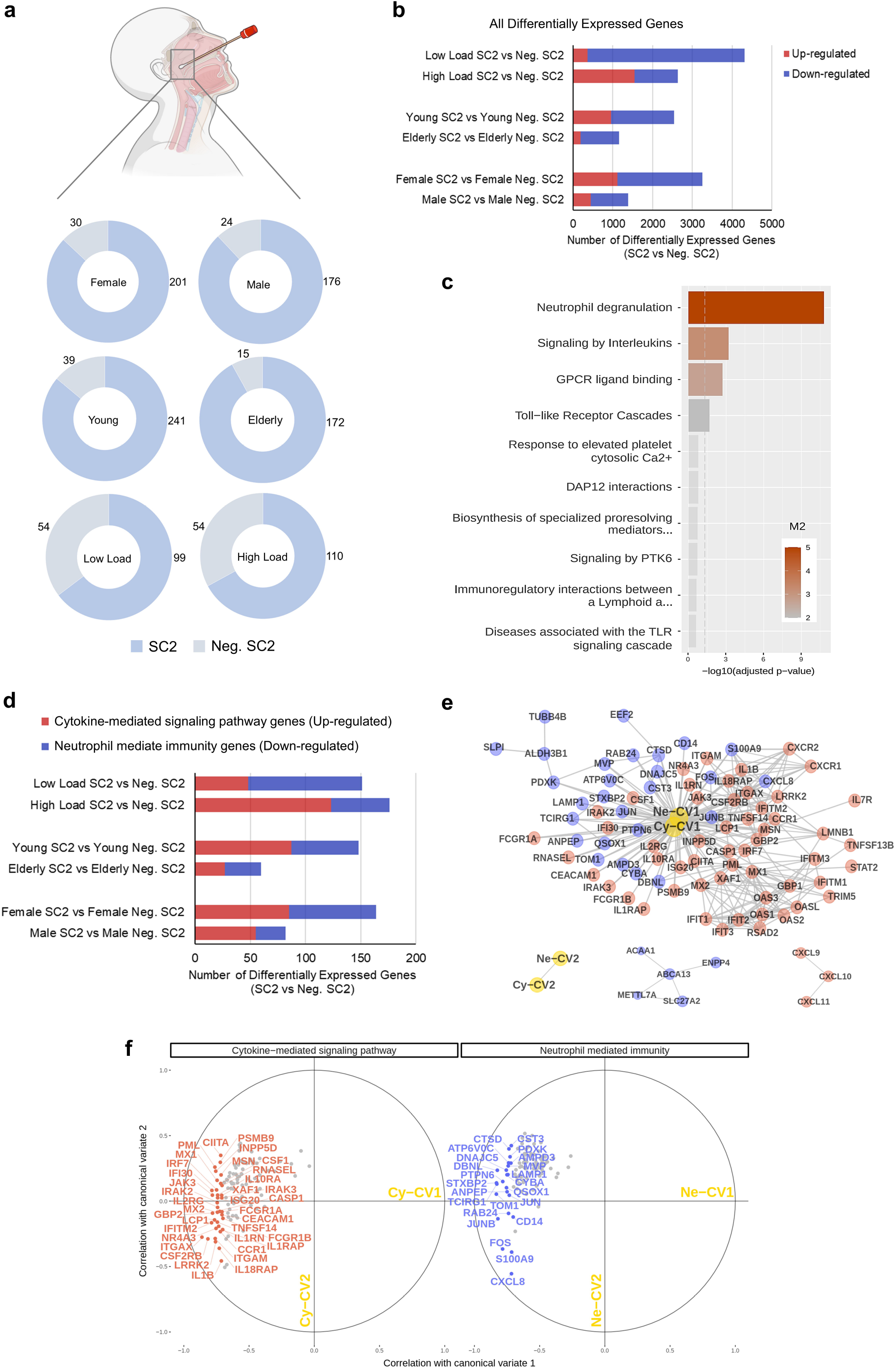
Transcriptomic analysis of swabs from SARS-CoV-2 positive compared with SARS-CoV-2 negative patients (dataset GSE152075). **a**, Number of SARS-CoV-2 (SC2) positive and negative samples by groups: gender, age (young < 60; elderly ≥ 60 years old) and viral load (low and high). **b**, The total number of differentially expressed genes in SC2 positive samples by group. **c**, Functional over-representation obtained by modular gene co-expression analysis (see Extended Data. Fig. 1) indicating a relationship between alterations in neutrophil activation and signaling by interleukins during the *in vivo* immune response to SARS-CoV-2. **d**, The total number of DEGs associated with cytokine-mediated signaling pathways (CMSP, red bars) and neutrophil-mediated immunity (NMI, blue bars) when comparing SC2 positive patients with corresponding viral load-, gender-, and age-matched negative SC2 samples. **e**, Heliographic and **f**, network representation of canonical correlation analysis of CMSP and NMI genes from SC2 positive patients supporting the correlation between these two gene ontology categories. CMSP and NMI genes with the Pearson correlation coefficient or *r* ≥ 0.7 are red and blue, respectively, while those with r < 0.7 are grey in both groups. Genes with corresponding principal component loadings < 0.7 have their names omitted. Cy-CV1: canonical variable 1 associated with CMSP genes; Cy-CV2: canonical variable 2 associated with CMSP genes; Ne-CV1: canonical variable 1 associated with NMI genes; Ne-CV2: canonical variable 2 associated with NMI genes.

Next, we evaluated the strength of the association between CMSP and NMI (described in **Supplementary Tables S6-S8**). Canonical-correlation analysis (CCA) revealed a strongly correlated network between these CMSP and NMI genes in the immune response to SARS-CoV-2 (**Fig. 1e, f**). Among others, the CCA showed an association between *IL-1β, IL-18* receptor accessory protein (*IL-18RAP*), C-C chemokine receptor type 1 (*CCR1*), interferon-induced guanylate-binding protein 2 (*GBP2*), interferon regulatory factor (*IRF7*) with NMI transcripts such as C-X-C motif chemokine ligand 8 (*CXCL8*, also called *IL-8*), lysosome-associated membrane protein 1 (*LAMP1*) and cytochrome B-245 Alpha Chain (*CYBA*). The association of these genes suggests an orchestrated modulation between interferon regulated genes (*GBP2, IRF7*, and *IFI30*), recruitment and neutrophil degranulation (*CXCL8* and *LAMP1*), and oxidative stress (*CYBA*). Taken together, the CCA analysis suggests that these two sets (CMSP and NMI) of highly correlated genes have a systemic representativity, suggesting pathophysiological relevance in COVID-19 patients.

### CMSP and NMI modular gene co-expression and enrichment in whole blood leukocytes from COVID-19 patients

Modular co-expression enrichment analyses of whole blood leukocytes from COVID-19 patients from a recent, publicly available dataset (GSE157103)^24^ showed coherent results compared to the swab dataset analyses. CEMITool analysis revealed ten co-expression modules enriched in whole leukocytes of COVID-19 patients compared to the control group. Among them, module M6 was enriched by interferon signaling genes, while module M7 was composed of genes associated with neutrophil degranulation in the COVID-19 group (**Fig. 2a-c**). The data, therefore, points toward a systemic (not restricted to upper airways) immunopathological association between interferon signaling and neutrophil-mediated immunity in patients with COVID-19. **Supplementary Tables S9** and **S10** describe the DEGs associated with CMSP and NMI in the GSE157103 dataset.

**Fig. 2.**
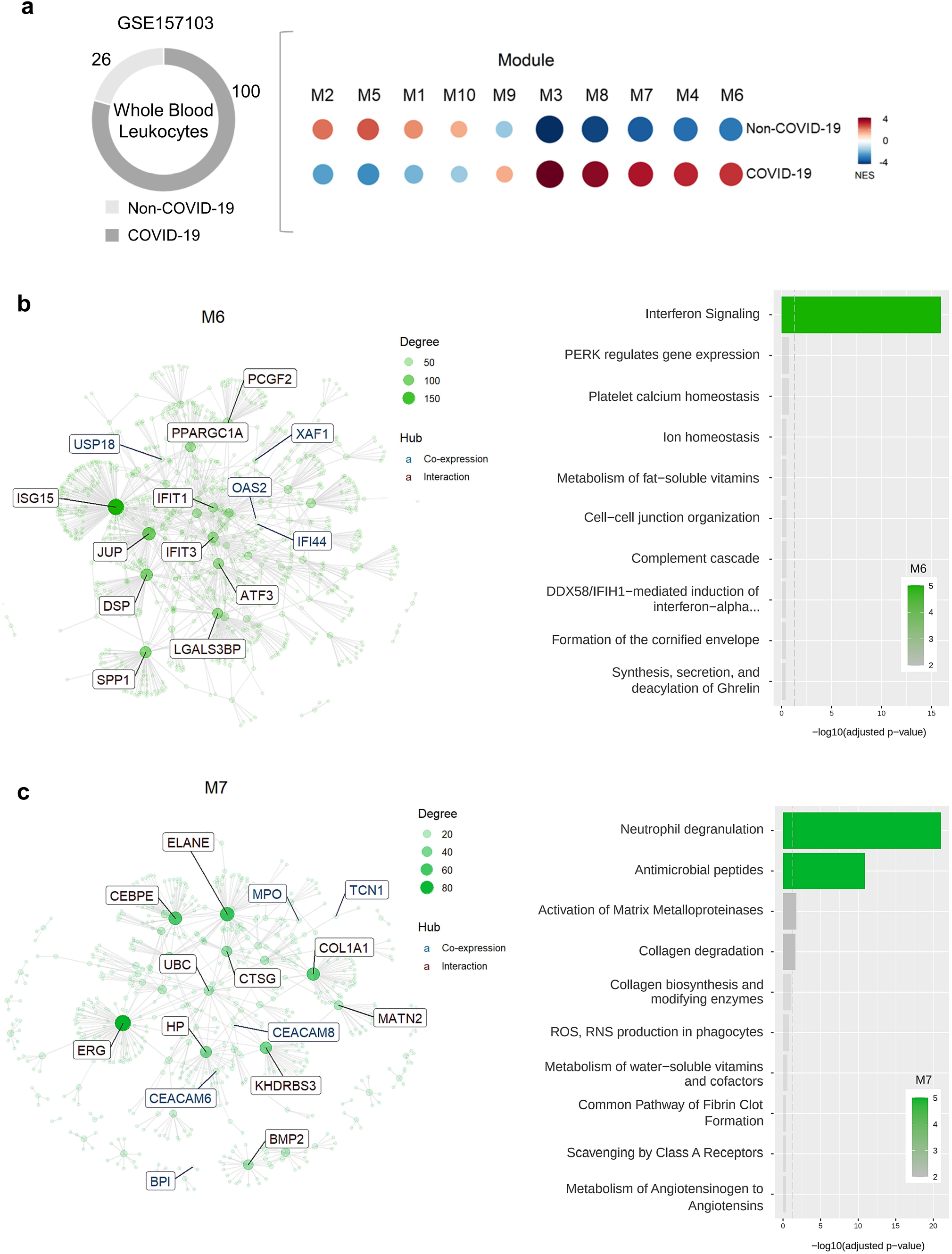
Modular gene co-expression analysis of differentially expressed genes of total leukocytes from SARS-CoV-2 patients (dataset GSE157103). **a**, Bubble heatmap showing the gene set enrichment of each module activity in peripheral blood leukocytes (PBLs) from SARS-CoV-2 (SC2) positive and negative subjects by gender, age (young < 60; elderly ≥ 60 years old), and severity (admitted or not at intensive care unit, ICU). Symbol size and color reflect the normalized enrichment score (NES). **b-d**, Functional over-representation interaction plot of gene co-expression modules, showing the enriched association between alterations in neutrophil degranulation and signaling by interleukins (module M1; **b**) as well as the enrichment of interferon signaling activation (module M4; **c**) and neutrophil degranulation (module M7; **d**) in PBLs. The interaction plots highlight gene nodes on each network, with potential hubs demonstrated inside rectangles and each node’s size proportionally to its degree.

### Effect of sex and age on cytokine-mediated signaling pathways in COVID-19 patients

To better understand the effect of gender and age on the anti-SARS-CoV-2 immune response in patients with COVID-19, we sought to further characterize the influence of gender and age on CMSP (detailed pro- or anti-inflammatory function of each gene is described in **Supplementary Table 11**). Multivariate comparison analysis of genes associated with CMSP identified in the nasopharyngeal swabs (GSE152075) revealed an upregulated spectrum of gene expression between SARS-CoV-2 positive (SC2) and negative (Neg. SC2) groups (**Fig. 3a**). Central tendency (median, **Fig. 3b**) and variability (interquartile range, **Extended Data Fig. 1**) analysis of CMSP transcriptional levels demonstrate the overall tendency in each SARS-CoV-2 positive and negative subgroups (female, male, young, elderly, high and low viral load). Hierarchical clustering analyses of gene expression revealed similarities between the CMSP signature of female and young groups and a close relationship between male and elderly patients. Female and young patients clustered with patients who had high viral loads, the latter presenting the patients with the most upregulated CMSP (**Fig. 3c and 3d)**. Conversely, males and elderly patients clustered near patients with low viral load. In this context, anti-viral transcripts induced by interferons^25^ were most upregulated in female, young and high-viral-load patients compared to male, elderly and low-viral-load patients. These included Interferon Induced Protein With Tetratricopeptide Repeats (*IFIT*)*1, IFIT2, IFIT3*; chemokine (C-X-C motif) ligand (*CXCL*)*9, CXCL10, CXCL11*; 2′, 5′-oligoadenylate synthetase (*OAS*)*1, OAS2*, and OAS3; tripartite motif (*TRIM*)*5* and *TRIM22*. The same phenomenon was observed in terms of transcripts of endogenous activators of the immune system, such as the *CD40* ligand and co-stimulatory molecules (*CD80* and *CD86*). Thus, our data suggest a higher transcriptional modulation of the immune response triggered by SARS-CoV-2 in female and young patients compared to males and elderly.

**Fig. 3.**
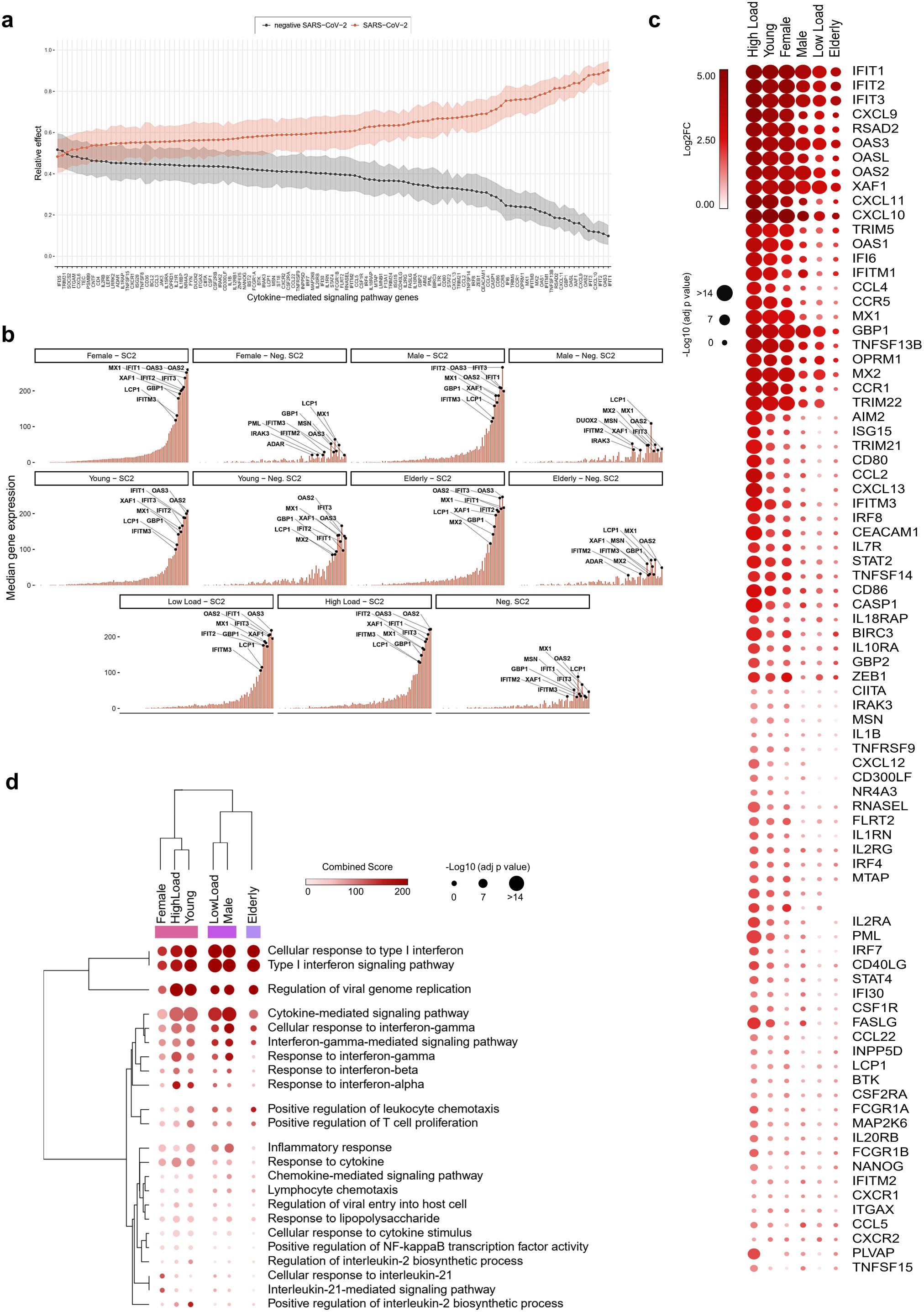
Cytokine-mediated signaling pathway signature by gender, age and viral load. **a**, Relative effects of transformed cytokine-mediated signaling pathway (CMSP) genes between SARS-CoV-2 positive (SC2) and negative (Neg. SC2) groups. Shaded areas indicate 95% percentile bootstrap confidence intervals for relative effects, based on 1000 bootstrap simulations. **b**, Central tendency (median) of gene expression for CMSP genes in each category versus the viral load-, gender-, and age-matched neg. SC2 controls. **c**, Bubble heatmap showing the expression pattern of CMSP genes by viral load, gender, and age groups. The size and color of circles correspond to -Log10 transformed adjusted *p*-value and Log2 fold change (Log2FC), respectively. The cutoff applied to identify the upregulated genes was Log2FC > 1 and adjusted *p*-value < 0.05. Rows and columns were clustered based on Euclidean distance between Log2FC values. **d**, Bubble heatmap representing the top-ranked combined scores for Biological Process (Gene Ontology) associated with CMSP genes in SC2 versus Neg. SC2 samples by groups. The circles’ size and color correspond to -Log10 transformed adjusted *P*-value and Combined Score, respectively. Rows and columns were clustered based on Euclidean distance between Combined Score values.

### Sex and age differences in neutrophil-mediated immunity

Patients with SARS-CoV-2 infections have been reported to have a higher number of MDSCs ^16–18^. In agreement with Schulte-Schrepping et al. ^26^, our differential expression analysis revealed several downregulated genes related to NMI are associated with MDSCs. These include *CYBA, CXCL8, CSTB, JUN, FOS*, and *MIF* ^26^ (**Fig. 4a and 4b**), which are involved in neutrophil degranulation and activation oxidative stress, and antimicrobial peptides in SARS-CoV-2 positive individuals. The detailed function of each gene is described in **Supplementary Table 12**. Among them are cathepsins (CTSB, CTSD, and CTSH), migration inhibitory factor (MIF), heat shock proteins (heat shock 70 kDa protein 1A or HSPA1A; and HSPA1B), and cytochrome B-245 Alpha Chain (CYBA). Hierarchical clustering analysis of NMI genes demonstrated that low viral load, female, and young patients presented a more downregulated transcriptomic profile (**Fig. 4c**). This was seen to a lesser extent in male and elderly patients. The cluster analysis suggests that these expression patterns reflect a close functional relationship (**Fig. 4d**) between low load, female and young patients.

**Fig. 4.**
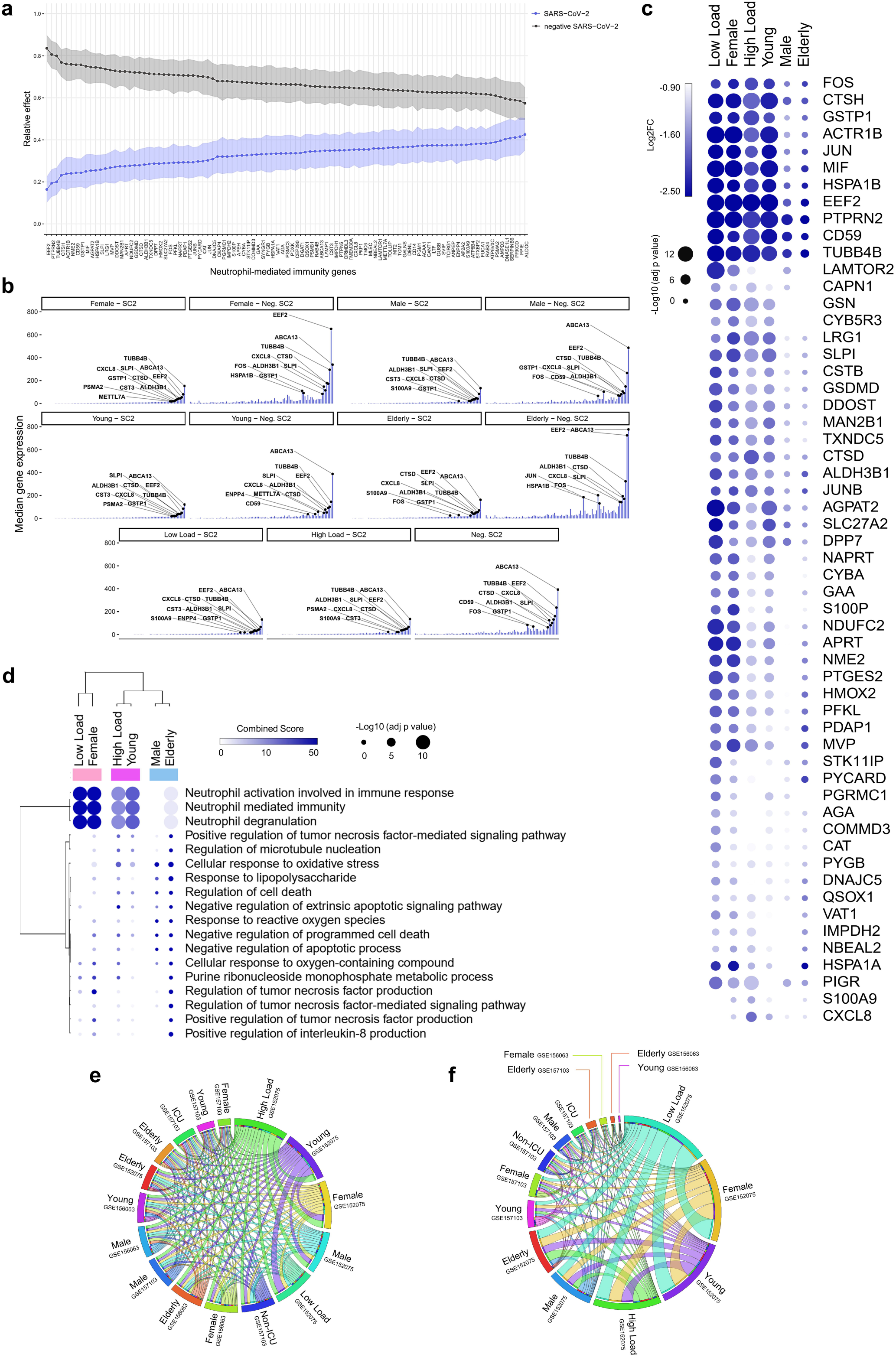
Neutrophil-mediated immunity signature by gender, age and viral load. **a**, Relative effects of transformed Neutrophil-mediated immunity (NMI) genes between SARS-CoV-2 positive (SC2) and negative (Neg. SC2) groups. Shaded areas indicate 95% percentile bootstrap confidence intervals for relative effects, based on 1000 bootstrap simulations. **b**, Central tendency (median) of gene expression for NMI genes in each category versus the viral load-, gender-, and age-matched neg. SC2 controls. **c**, Bubble heatmap showing the expression pattern of NMI genes by viral load, gender, and age groups. The size and color of circles correspond to -Log10 transformed adjusted *p*-value and Log2 fold change (Log2FC), respectively. The cutoff applied to identify the upregulated genes was Log2FC > 1 and adjusted *p*-value < 0.05. Rows and columns were clustered based on Euclidean distance between Log2FC values. **d**, Bubble heatmap representing the top-ranked combined scores for Biological Process (Gene Ontology) associated with NMI genes in SC2 versus Neg. SC2 samples by groups. The circles’ size and color correspond to -Log10 transformed adjusted *P*-value and Combined Score, respectively. Rows and columns were clustered based on Euclidean distance between Combined Score values. **e, f**, The thickness of each link in the Circos plot represents the number of shared cytokine-mediated signaling pathway (CMSP) (**e**) and NMI (**f**) genes throughout the datasets (GSE152075, GSE157103, and GSE156063) distributed by gender, age (young < 60; elderly ≥ 60 years old), viral load (low and high), and severity (admitted or not at intensive care unit, ICU).

We next analyzed the CMSP and NMI genes in two different datasets (GSE157103 and GSE156063). The circos plot showed that patients with a high viral load shared a higher number of CMSP genes with younger and female patients (**Fig. 4e**). Interestingly, female, young, and low-load patients shared more common NMI genes in swab and total leukocytes than male and elderly patients (**Fig. 4f**). This result suggests a predominant downregulation of NMI genes in groups of patients who do not develop severe disease. **Extended Data Fig. 2** shows the result of ridge regression analysis, which indicates associations of each gene belonging to the CMSP and NMI (described below) with viral load, in line with the formerly described results. Of note, several genes present in the dataset GSE152075 were also identified in another study of nasopharyngeal swabs developed by Mick et al. (**Extended Data Fig. 3**, dataset GSE156063), increasing the validity of these findings. Further, the upregulation of CMSP and NMI genes was present in both the datasets described above, which are of bulk RNAseq, and in a single-cell RNAseq study (**Extended Data Fig. 4**).

### Specific immune response-associated gene modulations in female infected with SARS-CoV-2

Considering the increased male/female mortality ratio (**Extended Data Fig. 5**), we sought to further characterize the transcriptomic features of COVID-19. We searched for sex differences between DEGs identified in female vs. male patients with COVID-19 as well as female vs. healthy male controls throughout the different RNAseq datasets. For instance, females with SARS-CoV2 showed reduced levels of *CXCL8* and its receptors (*CXCR1* and *CXCR2*) in different swab datasets (**Fig 5a-5d**). In addition, Mick’s and Liberman’s datasets presented a considerable overlap of DEGs and shared functional enrichment categories. We found that downregulated genes in females SC2 also enriched CMSP and NMI pathways (**Extended Data Fig. 6**). These shared genes are essential for conventional neutrophil chemotaxis and the recruitment of polymorphonuclear neutrophil (PMN)-MDSCs ^26^. Females also exhibited reduced levels of *IL-1β* when compared to male patients, a pro-inflammatory cytokine considered a potential target for COVID-19 therapy^27^. Other pro-inflammatory transcripts (neurobeachin like 2, *NBEAL2*) critical for leukocyte recruitment and granule exocytosis ^28,29^ or alarmins that are released during tissue damage (S100 Calcium Binding Protein A9, *S100A9*) ^30,31^ were lower in swabs from females infected with SARS-CoV-2 in comparison to males. Multivariate regression analysis indicated that the expression of genes shown in **Fig. 5a** and **5b** was strongly associated with the remaining genes belonging to CMSP and NMI (**Fig. 5g and Extended Data Figs. 7 and 8**). In addition, this approach indicates that females tend to systemically have a subtle lower gene expression pattern of immune genes compared to males infected with SARS-CoV-2.Likewise, **Extended Data Fig 7a** and **7b** show the multivariate regressions between these DEGs’ expression from females with the viral load and age, respectively. These results suggest that not all these genes demonstrate a divergent correlation by different viral loads and age, indicating that this modulation may be associated exclusively with male-female differences.

**Fig. 5.**
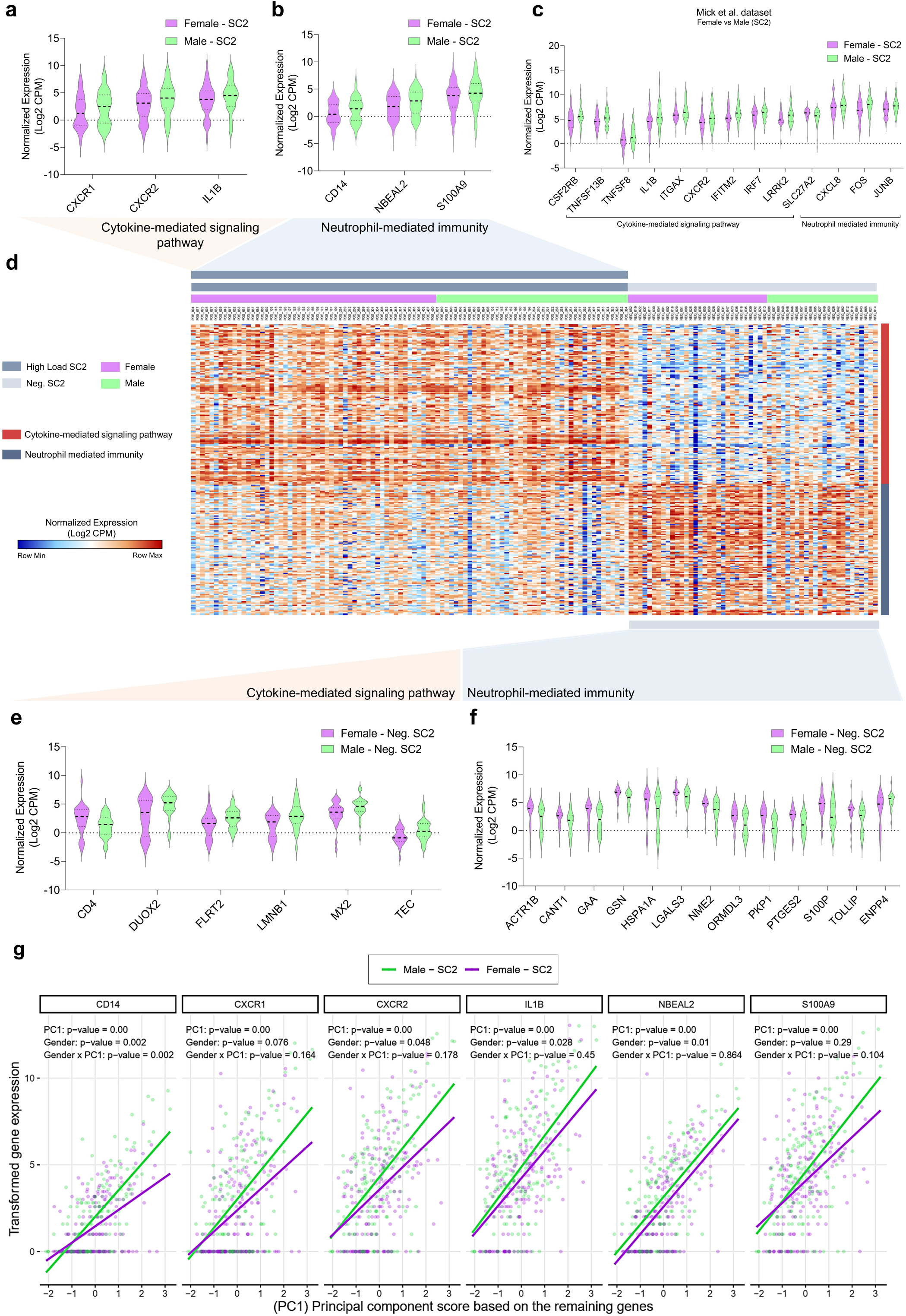
Transcriptional differences in cytokine-mediated signaling pathway and neutrophil-mediated immunity genes distinguish females and males. **a-b**, Violin plots present differentially expressed genes (DEGs, *p*-value < 0.05 and fold change (FC) >2) of cytokine-mediated signaling pathway (CMSP) (**a**) and neutrophil mediated immunity (NMI) (**b**) of SARS-CoV-2 positive (SC2) female samples versus SC2 male samples from the dataset GSE152075. **c**, Swab samples from positive SC2 patients from the dataset GSE156063 showing the DEGs associated with CMSP and NMI in female versus male samples. **d**, Unsupervised hierarchical clustering heatmap of CMSP and NMI genes (normalized gene expression in Log2 counts per million or CPM). On the right, red tracks denote CMSP genes, and blue tracks represent NMI genes. **e**, DEGs (*p*-value < 0.05 and FC >2) enriching cytokine-mediated signaling pathway in negative SC2 (Neg. SC2) female samples. **f**, DEGs enriching NMI of Neg. SC2 female versus Neg. SC2 male samples. All data of the violin plots are normalized expression (Log2 CPM). **g**, Multivariate regression modeling indicates that the expression values of the six genes (on the top, and also shown in **Fig. 5a** and **5b)** systemically associate with the expression of the other CMSP and NMI genes (shown in the X-axis and represented by the PC1 scores). Each dot represents a unique sample individual (purple: female; green: male). PC1: *p*-value < 0.05 indicates a significant association between the expression of 6 DEGs (vertical axis; *CD14, CXCR1, CXCR2, IL-1β, NBEAL2, S100A9*) with the PC1 score (expression of remaining genes, transformed by Log2(CPM+1), which are shown in the horizontal axis. Gender *p*-value ≤ 0.05 indicates significant differences in the mean expression of females compared to males regarding the 6 DEGs. Gender x PC1 *p*-value ≤ 0.05 indicates significant changes between females and males along the gradient (continuum) of the PC1 score.

Sex differences between DEGs of healthy controls showed downregulation of several genes belonging to CMSP compared to negative for SARS-CoV-2 males (**Fig 5e**). Among them are promoters of the H_2_O_2_ production in airways (NADPH oxidase enzymes Dual Oxidase 2, *DUOX2*)^32^, regulators of cell adhesion (Fibronectin Leucine-Rich Transmembrane protein 2, *FLRT2*), and proliferation (Laminin B-2). In contrast, NMI genes were upregulated in Neg. SC2 females compared to Neg. SC2 males (**Fig 5f**). These results show that even in the absence of the SARS-CoV-2, females present a different expression pattern of these genes, indicating that the immune response may be sex-dependent. This mucosal baseline state of reduced CMSP and increased NMI resembles an immune protective quiescent-like^33–35^ state. **Fig. 6a** summarizes these DEGs from females described above and their molecular function. Beyond the genes mentioned above, COVID-19 females presented seven exclusive DEGs (**Fig. 6b**) compared to the other SARS-CoV-2 positive groups (**Supplementary Table 8**). The Integrin Subunit Alpha M (*ITGAM*) participates in both IL-4/IL13 signaling and neutrophil degranulation, and three upregulated DEGs (*OPRD1, JAK3*, and *ITGAM*) is associated with IL-4/IL-13 signaling. *LRRK2* was not associated with any of these pathways. This signaling antagonizes the IFN-mediated immune response in the airways^36^. The other three downregulated DEGs (*JUP, DYNLL1*, and *DBNL*) belong to the neutrophil degranulation category. This differential expression reinforces the possibility of a better-modulated transcriptome profile of females in response to SARS-CoV-2.

In addition, to better understand this immune modulation profile underlying the relationship of CMSP and NMI gender dimorphism in COVID-19, we performed an Interferome analysis of these two sets of DEGs. This analysis revealed an interconnected network of IFN-regulated genes modulated either by IFN type I, II, and III (**Fig. 6b and Supplementary Tables 13 and 14**).

**Fig. 6.**
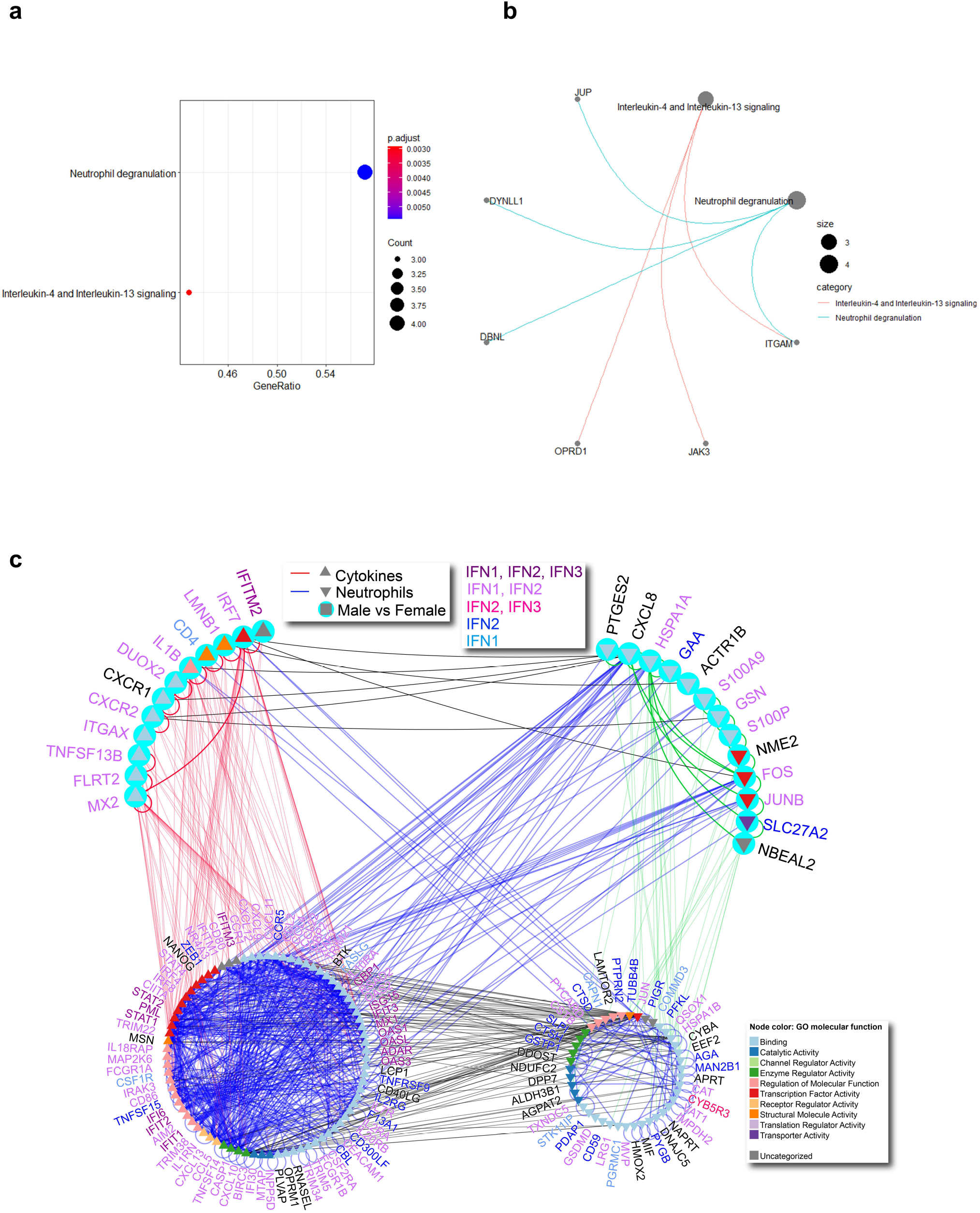
Network of cytokine-mediated signaling pathway and neutrophil-mediated immunity. **a**, Interactome between CMSP and NMI genes, highlighting the genes differentially expressed in female samples. Node color represents the GO molecular function associated with the gene. Triangles pointing up indicate CMSP genes, triangles pointing down indicate NMI genes. The light blue circles represent DEGs in females in both infected and uninfected samples. Blue edges highlight the NMI interactions; red edges reflect the CMSP interactions. The lower left subnetwork and lower right subnetwork show the interactions between CMSP and NMI genes, respectively. The label color represents the type of interferome associated with the gene. Interaction network was visualized using NAViGaTOR **b**, Dot plot presenting the two ontologies with adj. *p*-value < 0.01 enriched from exclusively female genes. X-axis presents the number of genes enriching the ontology term, and the dot size is proportional to this number. **c**, Interaction network of exclusive female genes. The node size is representing the number of genes according to each term.

## DISCUSSION

Here, we provide a systemic and integrative transcriptomic investigation aiming to identify signature changes that might explain the higher male/female mortality ratio observed in patients with COVID-19. We consistently found several upregulated CMSP and NMI DEGs throughout different public datasets of large patient cohorts infected with SARS-CoV-2. CCA and interferome analysis indicated that these DEGs form an interconnected network of several IFN-regulated genes. Females and young patients exhibited a higher capacity to trigger a higher transcriptional modulation than male and elderly patients. At the same time, females and young patients displayed downregulation of inflammatory genes such as CXCL8 receptors (*CXCR1* and *CXCR2*), *IL-1β, NBEAL2*, and *S100A9*. These genes are considered key players in immunological pathways involved in multiorgan injury and consequent death reported in COVID-19 ^16,17,26,37–39^. Our data is consistent with a recent report by Takahashi et *al*. showing lower plasma levels of *CXCL8* in females than male patients ^10^. Therefore, females may have protective transcriptional plasticity against harmful inflammation and the consequent tissue damage caused by the SARS-CoV-2 infection.

The role of neutrophils in the pathophysiology of COVID-19 is currently debated. While these cells are classically known to play an essential role in the immune response against bacterial and fungal infections, their antiviral function has only recently been characterized^40–42^. Neutrophils infiltrate the respiratory tract during viral infection and are required for a protective immune response against coronavirus. They also contribute significantly to respiratory tract pathology, i.e., hemorrhagic lesions, epithelial barrier permeability, and cellular inflammation in the lungs ^43^. Neutrophilia has consistently been found in COVID-19 patients and correlates with worse clinical outcomes^14,44–46^. However, recent studies assessing the activation status of neutrophils in COVID-19 patients have come to somewhat paradoxical results. Some investigators have identified that severe SARS-CoV-2 infection is associated with excessive release of reactive oxygen species (ROS) and neutrophil extracellular traps (NETs)^14,15^. Conversely, other studies demonstrated that the expansion of MDSCs increased with the severity of COVID-19, which agrees with our report of downregulated NMI signature ^16,18,47^.

MDSCs are a heterogeneous group of immature myeloid cells that have the capacity to suppress non-specific T and B lymphocyte responses ^18,48^. Much of our knowledge about MDSCs have been obtained through cancer studies ^47^, showing that tumors manipulate the myeloid system to evade the host immune response^49^. However, the physiological role of MDSCs has become increasingly evident in COVID-19. The results of several studies suggest that MDSCs are components of the healthy immune system and play a protective role in homeostatic and disease contexts. MDSCs expand when necessary to protect the host against tissue damage during autoimmune and inflammatory diseases^50,51^, traumatic stress, transplantation, and sepsis ^48,49,52^. While MDSCs may potentially represent biomarkers of COVID-19 severity, their presence suggests an attempt of the host to modulate the severe immune dysregulation triggered by SARS-CoV-2. In the same way, the high capacity of women to better modulate CMSP and NMI could represent a protective mechanism against severe COVID-19.

Notably, although playing a central role in the anti-viral immunity, hyper induction of IFNs following systemic activation of IFN-related genes have life-threatening immunopathological effects in COVID-19 ^37^. The interferome network, when well-orchestrated, is protective not only by promoting an anti-viral milieu but also by limiting airway inflammation by directly modulating pathogenic neutrophil accumulation^53^. Therefore, to address the threshold that shifts the IFN milieu from a protective to detrimental state, it will be imperative to identify markers that allow for appropriate therapeutic administration of IFN or JAK inhibitors to selected patients with COVID-19 ^54,55^.

While our study offers an explanation for the differences between mortality and morbidity between males and females with COVID-19, further potential mechanisms should be addressed. For instance, estrogen receptor signaling may protect against another coronavirus (SARS-CoV) ^56^. Furthermore, the sex-driven dimorphic immune response could be due to the location of several immune genes or immune regulatory genes on the X chromosome ^56^. Moreover, future studies need to evaluate the possible impact of sex-related risk factors for mortality in COVID-19 (smoking, alcohol consumption, and comorbidities) ^57–59^ on the immune response against SARS-CoV-2.

In conclusion, our integrative approach indicates that the gene expression profile associated with CMSP and NMI has a distinct pattern according to sex and age in patients with COVID-19. Thus, suggesting the existence of specific immune-regulatory pathways underlying the sexual dimorphism of COVID-19 morbidity and mortality. The specific profile of immune genes may help the future development of better targeted therapies to improve the outcomes of COVID-19.

## Online Methods

### Data Collection and Differential Expression Analysis

Lieberman et al., 2020^19^, and Mick et al., 2020^20^, recently performed RNASeq experiments investigating global transcriptional profiles of nasopharyngeal swabs from 668 individuals with SARS-CoV-2 (SC2) and 157 individuals negative of SARS-CoV-2 (Neg. SC2) (data accessible at NCBI GEO database^60^, accession number GSE GSE152075^19^ and GSE156063^20^). We retrieved the datasets to characterize the immunological signature. Read counts were transformed (log2 count per million or CPM), and differentially expressed transcripts between groups were identified through the webtool NetworkAnalyst 3.0 (https://www.networkanalyst.ca/)^61^ using limma-voom pipeline^62^. Age (< 60 young and ≥ 60 elderly individuals), gender (male and female), and viral load (only for GSE152075) categorized as previously described^19^. The cycle threshold (Ct) of the SARS-CoV-2 nucleocapsid gene region 1 (N1) target during diagnostic PCR: low viral load (N1 Ct > 24) and high viral load (N1 Ct < 19), defined the viral load. We applied the statistical cut-offs of log2 fold-change > 1 and adjusted p-value < 0.05 to determine DEGs between the categories. For subsequent analysis, we followed the limma-voom pipeline to identify DEGs between female and male.

### Enrichment Analysis and Data Visualization

We used these DEGs to identify different ontology terms. Biological processes [Gene Ontology (GO)] were analyzed using EnrichR (available at http://amp.pharm.mssm.edu/Enrichr/)^21,63^, and the enriched immunological terms were generated according to adjusted p-value < 0.05 and Z-score (correction to the test) in a combined score provided by EnrichR database^21,63^. The Biological Process terms were included in an integrative analysis using the criterion of over-representation (Log2 combined score > 2) in at least two categories. Concomitantly, we performed the analysis of gene co-expression modules with the R-package CEMiTool using default parameters^23^. We plotted the set of genes associated with CMSP (GO:0019221) and NMI (GO:0002446) in bubble-based heat maps with hierarchical clustering using web tool Morpheus (https://software.broadinstitute.org/morpheus/)^64^ with Euclidian distance metric. GraphPad Prisma v.8 (GraphPad Software) was used to generate the violin plots. The UniProtKB database (available at http://www.uniprot.org/) was used to access the functional information. We used clusterProfiler^65^ to obtain dot plots and Cnetplots of enriched terms associated with CMSP and NMI-associated genes. Shared CMSP and NMI genes among all groups were displayed using Circos Plot (http://circos.ca/). The identification of interferome genes was performed with Interferome V2.01 (http://www.interferome.org/interferome/home.jspx).

### Molecular Network of CMSP and NMI Genes

Network of cytokine-mediated signaling pathway and neutrophil-mediated immunity was constricted using DEGs, and highlighting the genes differentially expressed in female samples. DEGs were used as input into Integrated Interactions Database (IID version 2020-05; http://ophid.utoronto.ca/iid;^66,67^ to identify direct physical protein interactions. The resulting network was annotated, analyzed, and visualized using NAViGaTOR 3.013 (PMID: 19837718). Final network was exported in SVG format and finalized with legends in Adobe Illustrator. Node color represents GO molecular function as per legend. Triangles pointing up indicate CMSP genes, triangles pointing down indicate NMI genes. The light blue circles represent DEGs in females in both infected and uninfected samples. Blue edges highlight NMI interactions; red edges reflect CMSP interactions. The lower left subnetwork and lower right subnetwork show the interactions between CMSP and NMI genes, respectively. Protein name color represents the type of interferome associated with the gene.

### Statistics

Prior to the application of the statistical methods described below, the variable transformation was performed as described in each figure legend. For gene expression data, we added unity to all counts and consecutively applied a base two logarithmic function for each gene variable, herein called transformed gene expression. The remaining quantitative variables were scaled. Only CMSP and NMI genes were included in the data analysis as response variables. We used Nonparametric Multivariate Analysis of Variance (NP-MANOVA)^68^ to test differences on the mean vectors of gene expressions between SARS-CoV-2 positive (SC2) and negative (Neg. SC2) groups, separately for both CMSP and NMI genes.

#### Relative effect analysis

For each gene, relative effects^68^ of transformed expression were compared between SC2 and Neg. SC2 groups, with associated 95% confidence intervals, calculated via bootstrap simulation using the method of resampling pairs^69^. Bootstrap statistics were based on 1000 simulations and percentile confidence intervals^69^.

#### Canonical Correlation Analysis

Canonical Correlation Analysis (CCA)^70^ was applied in order to investigate patterns of association between genes related to CMSP and NMI genes, considering the observations from the SC2 group alone. We retained the first two canonical variates (CVs) for subsequent interpretations.

#### Ridge Regression

We used Ridge Regression^71^ for setting up a predictive model for the response variable viral load as a function of the regression covariates age, gender, and transformed gene expressions, considering observations from the SC2 group alone. Model estimates were obtained using 10-fold cross validation^71^. As a testing set, 25% of the dataset was kept and used for evaluating the model prediction accuracy.

#### Multivariate Regression

We performed Multivariate Regression (MVR) analysis with normally distributed additive errors^72^ to model the mean vector of transformed gene expression associated with the six genes found with differentially expression in females (CD14, CXCR1, CXCR2, IL1B, NBEAL2, and S100A9), considering the observations from the SC2 group alone. The regression covariates included were the principal component scores of the remaining genes and variables age and viral load. These scores resulted from a Principal Component Analysis (PCA) based on the transformed gene expressions for genes not included as responses in the MVR mode. After PCA, the estimated matrix of loadings was rotated using the varimax criterion. The Likelihood Ratio (LR) and Wald statistics were used, respectively, for testing the generalized hypothesis involving MRV model parameters and obtaining the statistical significance of each regression slope individually^72^. The level of significance for all hypothesis testing was fixed at 5%, and the wild bootstrap method was used for calculating p-values and parameter’s standard errors robust against heteroscedasticity on the regression errors^73^. Bootstrap statistics were based on 1000 simulations^69^. Model adequacy was studied using the metrics developed in Díaz-García et al., 2003^72^.

### Statistical Software and packages

The sample median and sample interquartile range were calculated using R software version 4.0.2 (https://www.r-project.org/index.html). The NP-MANOVA, CCA, MVR, and Ridge Regression analysis were all performed on the R software version 4.0.2. Specifically, for NP-MANOVA, it was used the npmv package^68^ for CCA the whitening^70^, DFA, and CANCOR packages, for Ridge Regression the glmnet package and for PCA the psych package. MVR was implemented by the authors following the results of García et al. (2003)^72^. Finally, all statistical graphs were constructed using the functionalities of the ggplot2 package^74^.

### Ethical Statement

Since we used publicly available data Ethical approval was not applicable.

## Data Availability

The published bulk RNAseq datasets from patients can be found in the GEO database (GSE152075, GSE157103, GSE156063). Single-cell RNAseq data (dataset EGA00001004571) was obtained from neutrophil clusters, as reported by Schulte-Schrepping et al.

## Acknowledgments

We acknowledge the São Paulo Research Foundation (FAPESP grants 2017/05264-7 to NOSC, 2018/18886-9 to OCM, 2020/01688-0 to OCM, 2020/07069-0 to OCM, 2020/07972-1 to OTC, and 2020/09146-1 to PPF) for financial support. Computational analysis was supported in part by the grants from Ontario Research Fund (#34876), Natural Sciences Research Council (NSERC #203475), Canada Foundation for Innovation (CFI #29272, #225404, #33536) and IBM. This study was financed in part by the Coordenação de Aperfeiçoamento de Pessoal de Nível Superior - Brasil (CAPES) - Finance Code 001.

## Author Contributions

PPF, AM, OCM co-wrote the manuscript and provided scientific insights; PPF, AM, TDCH, IJ, and OCM, performed bioinformatics analyses; PPF, GCB, RFC, LCSF, ACN, HN, NOSC, VC, and OCM conceived and designed the study; PPF, AM, GCB, ISF, LFS, RCS, SN, DRP, GCM, OCM, analyzed data; PPF, AM, GLM, LFS, NEK, GLM, GCM, RFC, LCS, ACN, HN, IJ, HDO, NOSC, VC, and OCM revised and edited the final manuscript; VC and OCM supervised the project.

## Competing interest statement

The authors declare no competing financial and/or non-financial interests in relation to the work described.

## Data availability

The published bulk RNAseq datasets can be found in the GEO database (GSE152075, GSE157103, GSE156063). Single-cell RNAseq data **(dataset EGA00001004571)** was obtained from neutrophil clusters, as reported by Schulte-Schrepping et al^75^.

---

**Extended Data Fig. 1| Interquartile range of gene expression by gender, age, and viral load. a-b**, Measure of variability (interquartile range) of gene expressions for (**a**) cytokine-mediated signaling pathway (CMSP) genes and (**b**) neutrophil-mediated immunity (NMI) genes by categories viral load-, gender-, and age-matched SARS-CoV-2 (SC2) positive vs. Negative (Neg. SC2).

**Extended Data Fig. 2| The relationship between gene expression and viral load. a**, The association between the expression of CMSP and NMI genes and viral load was calculated using Ridge regression estimates as described in the material and methods. Due to the N1.Ct1 values (viral load) being inversely proportional to the amount of viruses detected by PCR, genes with higher regression coefficient estimate tends to correlate negatively with the viral load, and conversely. Only observations from the positive SC2 group were used for the estimation of parameters in the model. CMSP: cytokine-mediated signaling pathway, NMI: neutrophil mediated-immunity. SC2 = SARS-CoV-2 positive. **b**, Accuracy of the ridge regression model fitted for estimating viral load as a linear function of age, gender and transformed gene expressions. Each dot corresponds to an individual observation of the testing dataset, which was composed of 25% of the sample (total SC2 positive group).

**Extended Data Fig. 3| Validation of cytokine-mediated signaling pathway and neutrophil-mediated immunity genes in swab samples (GSE156063). a, c, e, g**, Upsetplot displaying the set intersections for upregulated and downregulated genes in the Lieberman *et al*. dataset (GSE152075) and Mick *et al*. dataset (GSE156063) female samples (**a**), male (**c**), young (**e**), and elderly (**g**). **b, d, f, h**, Boxplot showing the common differentially expressed genes in female samples (**b**), male (**d**), young (**f**), and elderly (**h**) of the Mick et al. dataset. Data are presented as normalized expression (Log2 CPM) SARS-CoV-2 vs. negative SC2 samples, considering only the cytokine-mediated signaling pathway (CMSP) and neutrophil-mediated immunity (NMI) genes with adj *p*-value < 0.05 and Log2FC >1. Set 1: upregulated in Lieberman *et al*. dataset (GSE152075); Set 2: downregulated in Lieberman *et al*. dataset (GSE152075); Set 3: upregulated in Mick *et al*. dataset (GSE156063); Set 4: downregulated in Mick *et al*. dataset (GSE156063).

**Extended Data Fig. 4| Upregulated CMSP genes and downregulated NMI genes idenfied by single cell RNAseq (dataset EGAS00001004571) a**,**b**, Dot plot of enriched terms **(a)** upregulated and (**b**) downregulated in neutrophil clusters obtained as described by Schulte-Schrepping et al^86^. **c, d**, Cnetplot of CMSP and NMI-associated genes. Graphics were obtained using clusterProfiler.

**Extended Data Fig. 5| Percentage of deaths due to COVID-19 is higher in males than in females. a**, Percentage of male and female deaths by countries. Data were retrieved from https://globalhealth5050.org/the-sex-gender-and-covid-19-project/dataset/.

**Extended Data Fig. 6| Enrichment analysis of downregulated genes in female samples compared with male samples from the Mick et al. dataset (GSE156063). a**, Most enriched biological process terms (adj *p*-value< 0.05) with downregulated genes in female samples positive for SARS-CoV-2 compared with male samples positive for SARS-CoV-2 (validation dataset GSE156063). **b**, most enriched biological process terms (adj *p*-value< 0.05) with downregulated genes in female samples negative for SARS-CoV-2 compared with male samples negative for SARS-CoV-2 (validation dataset GSE156063).

**Extended Data Fig. 7| Relationship between the gene expression with the viral load and age. a-b**, Multivariate regression modeling indicates the expression values of the 6 genes (*CD14, CXCR1, CXCR2, IL-1*β, *NBEAL2, S100A9*) and their association with **a**, Viral load and **b**, Age. Each dot represents a unique individual sample (purple: female; green: male). Viral load or Age: *p*-value < 0.05 indicates a significant association of these variables with the expression of these 6 DEGs (transformed by Log2(CPM+1), which are shown in the horizontal axis. Gender *p*-value ≤ 0.05 indicates significant differences in the mean expression of females compared to males regarding the 6 DEGs. Gender x Viral load or Age: *p*-value ≤ 0.05 indicates significant changes between female and male along the gradient (continuum) of viral load or age. Scaled means that the variable was transformed by subtracting its mean and division by its standard deviation.

**Extended Data Fig. 8| Relationship between the profile of gene expression respect to viral load and age. a**, Network showing the correlation between genes and its association with the principal component (PC) analysis performed for obtaining the regression covariate PC1 included in the multivariate regression model described in the Statistical Methods section. Only observations from the SC2 group were used for analysis. CMSP and NMI genes are colored in red and blue, respectively. Pearson correlations ≥ 0.7 are indicated by grey lines. The size of nodes and gene names are directly proportional to the principal component loadings. Genes with corresponding principal component loadings < 0.7 have their names omitted. The loading estimates were all positive after varimax rotation. **b**, Graphs for residual analysis and diagnostics of the multivariate regression model fitted for modeling the transformed gene expression of *CD14, CXCR1, CXCR2, IL1*β, *NBEAL2*, and *S100A9* as a linear function of the covariates age, gender, viral load (n1_ct), and PC1. For the Gender x Index of the observation graph, the value 0 indicates male, and the value 1 indicates female. PC1: scores for each observation on the first principal component based on the remaining genes not included as response variables in the model.

## Notes

### Competing Interest Statement

The authors have declared no competing interest.

### Clinical Trial

Since we used publicly available data Ethical approval or trial ID was not applicable. The published bulk RNAseq datasets from patients can be found in the GEO database (GSE152075, GSE157103, GSE156063).

### Funding Statement

We acknowledge the Sao Paulo Research Foundation (FAPESP grants 2017/05264-7 to NOSC, 2018/18886-9 to OCM, 2020/01688-0 to OCM, 2020/07069-0 to OCM, 2020/07972-1 to OTC, and 2020/09146-1 to PPF) for financial support. Computational analysis was supported in part by the grants from Ontario Research Fund (#34876), Natural Sciences Research Council (NSERC #203475), Canada Foundation for Innovation (CFI #29272, #225404, #33536) and IBM. This study was financed in part by the Coordenacao de Aperfeicoamento de Pessoal de Nivel Superior - Brasil (CAPES) - Finance Code 001.

### Author Declarations

Since we used publicly available data Ethical approval was not applicable.

## REFERENCES

1. Li, R. et al. Substantial undocumented infection facilitates the rapid dissemination of novel coronavirus (SARS-CoV-2). Science (80-.). 368, 489–493 (2020).

2. Lescure, F. X. et al. Clinical and virological data of the first cases of COVID-19 in Europe: a case series. Lancet Infect. Dis. 20, 697–706 (2020).

3. Progress report on the coronavirus pandemic. Nature 584, 325 (2020).

4. WHO Coronavirus Disease (COVID-19) Dashboard | WHO Coronavirus Disease (COVID-19) Dashboard.

5. Sun, P., Qie, S., Liu, Z., Ren, J. & Xi, J. Clinical Characteristics of 5732 Patients with 2019-nCoV Infection. SSRN Electron. J. (2020) doi:10.2139/ssrn.3539664.

6. Song, J. W. et al. Immunological and inflammatory profiles in mild and severe cases of COVID-19. Nat. Commun. 11, 1–10 (2020).

7. Hoffmann, M. et al. SARS-CoV-2 Cell Entry Depends on ACE2 and TMPRSS2 and Is Blocked by a Clinically Proven Protease Inhibitor. Cell 181, 271-280.e8 (2020).

8. Bhopal, S. S. & Bhopal, R. Sex differential in COVID-19 mortality varies markedly by age. The Lancet vol. 396 532–533 (2020).

9. Sakuishi, K. et al. Targeting Tim-3 and PD-1 pathways to reverse T cell exhaustion and restore anti-tumor immunity. J. Exp. Med. (2010) doi:10.1084/jem.20100643.

10. Takahashi, T. et al. Sex differences in immune responses that underlie COVID-19 disease outcomes. Nature 1–6 (2020) doi:10.1038/s41586-020-2700-3.

11. Chen, Z. & John Wherry, E. T cell responses in patients with COVID-19. Nature Reviews Immunology vol. 20 529–536 (2020).

12. Merad, M. & Martin, J. C. Pathological inflammation in patients with COVID-19: a key role for monocytes and macrophages. Nature Reviews Immunology vol. 20 355–362 (2020).

13. Moore, J. B. & June, C. H. Cytokine release syndrome in severe COVID-19. Science vol. 368 473–474 (2020).

14. Laforge, M. et al. Tissue damage from neutrophil-induced oxidative stress in COVID-19. Nature Reviews Immunology vol. 20 515–516 (2020).

15. Zuo, Y. et al. Neutrophil extracellular traps in COVID-19. JCI Insight 5, (2020).

16. Schulte-Schrepping, J. et al. Severe COVID-19 Is Marked by a Dysregulated Myeloid Cell Compartment. Cell (2020) doi:10.1016/j.cell.2020.08.001.

17. Silvin, A. et al. Elevated Calprotectin and Abnormal Myeloid Cell Subsets Discriminate Severe from Mild COVID-19. Cell 182, 1401 (2020).

18. Agrati, C. et al. Expansion of myeloid-derived suppressor cells in patients with severe coronavirus disease (COVID-19). Cell Death Differ. 1–12 (2020) doi:10.1038/s41418-020-0572-6.

19. Lieberman, N. A. P. et al. In vivo antiviral host transcriptional response to SARS-CoV-2 by viral load, sex, and age. PLOS Biol. 18, e3000849 (2020).

20. Mick, E. et al. Upper airway gene expression differentiates COVID-19 from other acute respiratory illnesses and reveals suppression of innate immune responses by SARS-CoV-2. medRxiv Prepr. Serv. Heal. Sci. (2020) doi:10.1101/2020.05.18.20105171.

21. Kuleshov, M. V et al. Enrichr: a comprehensive gene set enrichment analysis web server 2016 update. Nucleic Acids Res. 44, W90–7 (2016).

22. Chen, E. Y. et al. Enrichr: interactive and collaborative HTML5 gene list enrichment analysis tool. BMC Bioinformatics 14, 128 (2013).

23. Russo, P. S. T. et al. CEMiTool: a Bioconductor package for performing comprehensive modular co-expression analyses. BMC Bioinformatics 19, 56 (2018).

24. Overmyer, K. A. et al. Large-Scale Multi-omic Analysis of COVID-19 Severity. Cell Syst. (2020) doi:10.1016/j.cels.2020.10.003.

25. Rusinova, I. et al. Interferome v2.0: an updated database of annotated interferon-regulated genes. Nucleic Acids Res. 41, D1040–6 (2013).

26. Sun, L. et al. Inhibiting myeloid-derived suppressor cell trafficking enhances T cell immunotherapy. JCI Insight 4, (2019).

27. Huet, T. et al. Anakinra for severe forms of COVID-19: a cohort study. Lancet Rheumatol. (2020) doi:10.1016/S2665-9913(20)30164-8.

28. Sowerby, J. M. et al. NBEAL2 is required for neutrophil and NK cell function and pathogen defense. J. Clin. Invest. 127, 3521–3526 (2017).

29. Tariket, S., Guerrero, J. A., Garraud, O., Ghevaert, C. & Cognasse, F. Platelet α-granules modulate the inflammatory response under systemic lipopolysaccharide injection in mice. Transfusion 59, 32–38 (2019).

30. Crowe, L. A. N. et al. S100A8 & S100A9: Alarmin mediated inflammation in tendinopathy. Sci. Rep. 9, 1–12 (2019).

31. Wang, S. et al. S100A8/A9 in inflammation. Frontiers in Immunology vol. 9 1298 (2018).

32. Fink, K. et al. IFNβ/TNFα synergism induces a non-canonical STAT2/IRF9-dependent pathway triggering a novel DUOX2 NADPH Oxidase-mediated airway antiviral response. Cell Res. 23, 673–690 (2013).

33. Card, C. M., Ball, T. B. & Fowke, K. R. Immune Quiescence: A model of protection against HIV infection. Retrovirology vol. 10 141 (2013).

34. Yao, X. D. et al. Acting locally: Innate mucosal immunity in resistance to HIV-1 infection in Kenyan commercial sex workers. Mucosal Immunol. 7, 268–279 (2014).

35. McLaren, P. J. et al. HIV-exposed seronegative commercial sex workers show a quiescent phenotype in the CD4+ T cell compartment and reduced expression of HIV-dependent host factors. in Journal of Infectious Diseases vol. 202 (J Infect Dis, 2010).

36. Zissler, U. M. et al. Interleukin-4 and interferon-γ orchestrate an epithelial polarization in the airways. Mucosal Immunol. 9, 917–926 (2016).

37. Broggi, A. et al. Type III interferons disrupt the lung epithelial barrier upon viral recognition. Science 369, 706–712 (2020).

38. Jose, R. J. & Manuel, A. COVID-19 cytokine storm: the interplay between inflammation and coagulation. The Lancet Respiratory Medicine vol. 8 e46–e47 (2020).

39. Condamine, T., Mastio, J. & Gabrilovich, D. I. Transcriptional regulation of myeloid-derived suppressor cells. J. Leukoc. Biol. 98, 913–922 (2015).

40. Gabriel, C., Her, Z. & Ng, L. F. P. Neutrophils: Neglected players in viral diseases. DNA and Cell Biology vol. 32 665–675 (2013).

41. Naumenko, V., Turk, M., Jenne, C. N. & Kim, S. J. Neutrophils in viral infection. Cell and Tissue Research vol. 371 505–516 (2018).

42. Galani, I. E. & Andreakos, E. Neutrophils in viral infections: Current concepts and caveats. J. Leukoc. Biol. 98, 557–564 (2015).

43. Haick, A. K., Rzepka, J. P., Brandon, E., Balemba, O. B. & Miura, T. A. Neutrophils are needed for an effective immune response against pulmonary rat coronavirus infection, but also contribute to pathology. J. Gen. Virol. 95, 578–590 (2014).

44. Zhou, Z. et al. Heightened Innate Immune Responses in the Respiratory Tract of COVID-19 Patients. Cell Host Microbe 27, 883-890.e2 (2020).

45. Huang, C. et al. Clinical features of patients infected with 2019 novel coronavirus in Wuhan, China. Lancet 395, 497–506 (2020).

46. Giamarellos-Bourboulis, E. J. et al. Complex Immune Dysregulation in COVID-19 Patients with Severe Respiratory Failure. Cell Host Microbe 27, 992-1000.e3 (2020).

47. Kumar, V., Patel, S., Tcyganov, E. & Gabrilovich, D. I. The Nature of Myeloid-Derived Suppressor Cells in the Tumor Microenvironment. Trends in Immunology vol. 37 208–220 (2016).

48. Condamine, T. & Gabrilovich, D. I. Molecular mechanisms regulating myeloid-derived suppressor cell differentiation and function. Trends in Immunology vol. 32 19–25 (2011).

49. Gabrilovich, D. I., Ostrand-Rosenberg, S. & Bronte, V. Coordinated regulation of myeloid cells by tumours. Nature Reviews Immunology vol. 12 253–268 (2012).

50. Kollmann, T. R., Kampmann, B., Mazmanian, S. K., Marchant, A. & Levy, O. Protecting the Newborn and Young Infant from Infectious Diseases: Lessons from Immune Ontogeny. Immunity vol. 46 350–363 (2017).

51. Gabrilovich, D. I. & Nagaraj, S. Myeloid-derived suppressor cells as regulators of the immune system. Nature Reviews Immunology vol. 9 162–174 (2009).

52. Crook, K. R. Role of myeloid-derived suppressor cells in autoimmune disease. World J. Immunol. 4, 26 (2014).

53. Nandi, B. & Behar, S. M. Regulation of neutrophils by interferon-γ limits lung inflammation during tuberculosis infection. J. Exp. Med. 208, 2251–2262 (2011).

54. Lee, J. S. & Shin, E. C. The type I interferon response in COVID-19: implications for treatment. Nature Reviews Immunology 1–2 (2020) doi:10.1038/s41577-020-00429-3.

55. Acharya, D., Liu, G. Q. & Gack, M. U. Dysregulation of type I interferon responses in COVID-19. Nature Reviews Immunology vol. 20 397–398 (2020).

56. Scully, E. P., Haverfield, J., Ursin, R. L., Tannenbaum, C. & Klein, S. L. Considering how biological sex impacts immune responses and COVID-19 outcomes. Nat. Rev. Immunol. 20, 442–447 (2020).

57. Vardavas, C. I. & Nikitara, K. COVID-19 and smoking: A systematic review of the evidence. Tobacco Induced Diseases vol. 18 (2020).

58. Gagliardi, M. C., Tieri, P., Ortona, E. & Ruggieri, A. ACE2 expression and sex disparity in COVID-19. Cell Death Discovery vol. 6 1234567890 (2020).

59. Kopel, J. et al. Racial and Gender-Based Differences in COVID-19. Frontiers in Public Health vol. 8 418 (2020).

60. Edgar, R., Domrachev, M. & Lash, A. E. Gene Expression Omnibus: NCBI gene expression and hybridization array data repository. Nucleic Acids Res. 30, 207–210 (2002).

61. Zhou, G. et al. NetworkAnalyst 3.0: A visual analytics platform for comprehensive gene expression profiling and meta-analysis. Nucleic Acids Res. 47, W234–W241 (2019).

62. Law, C. W., Chen, Y., Shi, W. & Smyth, G. K. Voom: Precision weights unlock linear model analysis tools for RNA-seq read counts. Genome Biol. 15, R29 (2014).

63. Chen, E. Y. et al. Enrichr: interactive and collaborative HTML5 gene list enrichment analysis tool. BMC Bioinformatics 14, 128 (2013).

64. Starruß, J., de Back, W., Brusch, L. & Deutsch, A. Morpheus: a user-friendly modeling environment for multiscale and multicellular systems biology. Bioinformatics 30, 1331–2 (2014).

65. Yu, G., Wang, L. G., Han, Y. & He, Q. Y. ClusterProfiler: An R package for comparing biological themes among gene clusters. Omi. A J. Integr. Biol. 16, 284–287 (2012).

66. Kotlyar, M., Pastrello, C., Malik, Z. & Jurisica, I. IID 2018 update: Context-specific physical protein-protein interactions in human, model organisms and domesticated species. Nucleic Acids Res. 47, D581–D589 (2019).

67. Pastrello, C., Kotlyar, M. & Jurisica, I. Informed use of protein–protein interaction data: A focus on the integrated interactions database (IID). in Methods in Molecular Biology vol. 2074 125–134 (Humana Press Inc., 2020).

68. Ellis, A. R., Burchett, W. W., Harrar, S. W. & Bathke, A. C. Nonparametric inference for multivariate data: The R package npmv. J. Stat. Softw. 76, 1–18 (2017).

69. Eck, D. J. Bootstrapping for multivariate linear regression models. Stat. Probab. Lett. 134, 141–149 (2018).

70. Jendoubi, T. & Strimmer, K. A whitening approach to probabilistic canonical correlation analysis for omics data integration. BMC Bioinformatics 20, 15 (2019).

71. Engebretsen, S. & Bohlin, J. Statistical predictions with glmnet. Clin. Epigenetics 11, 123 (2019).

72. Díaz-García, J. A., Rojas, M. G. & Leiva-Sánchez, V. Influence diagnostics for elliptical multivariate linear regression models. Commun. Stat. - Theory Methods 32, 625–641 (2003).

73. Flachaire, E. Bootstrapping heteroskedasticity consistent covariance matrix estimator. Comput. Stat. 17, 501–506 (2002).

74. Wickham, H. Getting Started with ggplot2. in 11–31 (2016). doi:10.1007/978-3-319-24277-4_2.

75. Schulte-Schrepping, J. et al. Severe COVID-19 Is Marked by a Dysregulated Myeloid Cell Compartment. Cell 182, 1419-1440.e23 (2020).

