## Supplementary figures and images for "Specific immune-regulatory transcriptional signatures reveal sex and age differences in SARS-CoV-2 infected patients"

### ExtendedDataFig.1.tiff

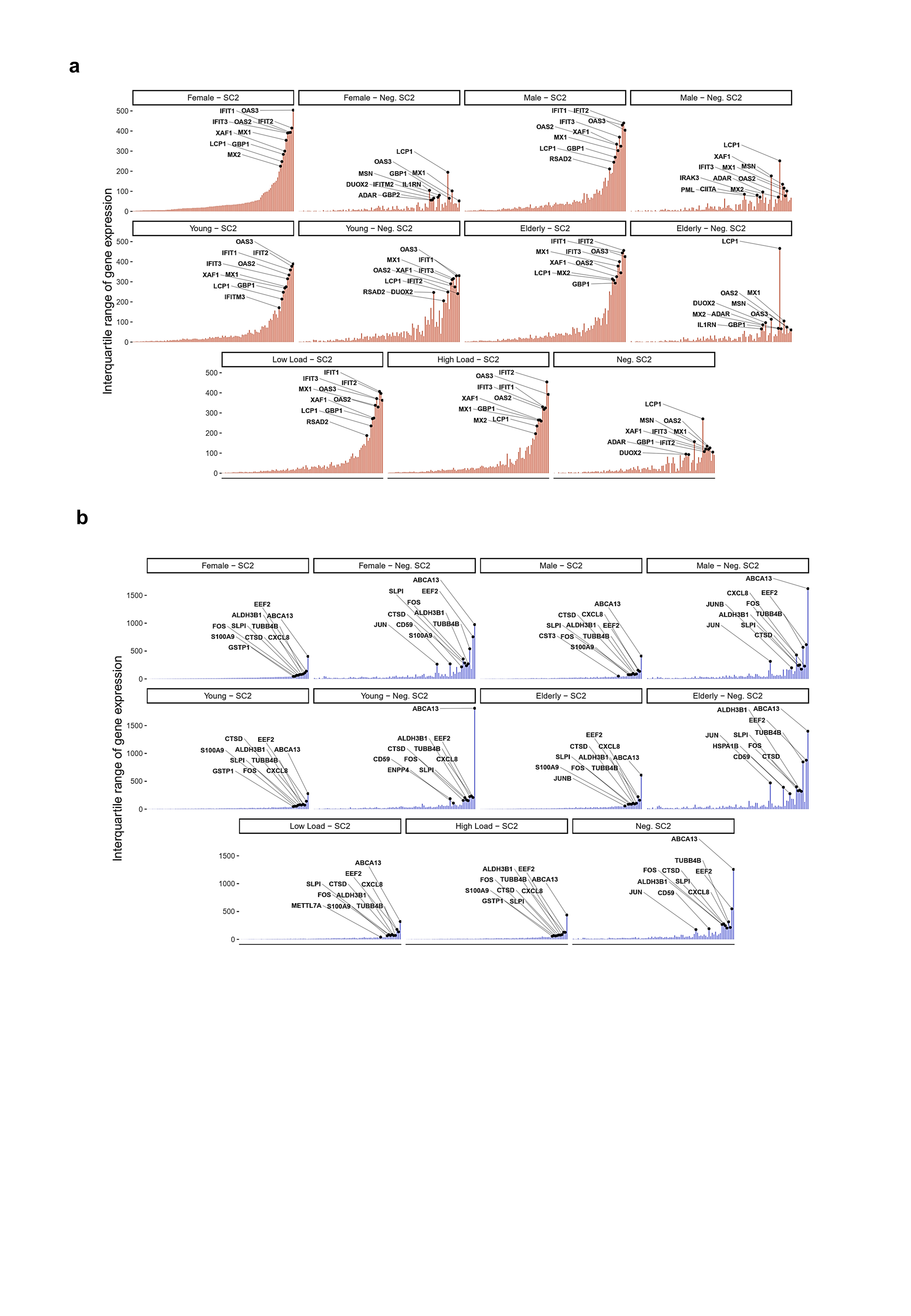

### ExtendedDataFig.2.tiff

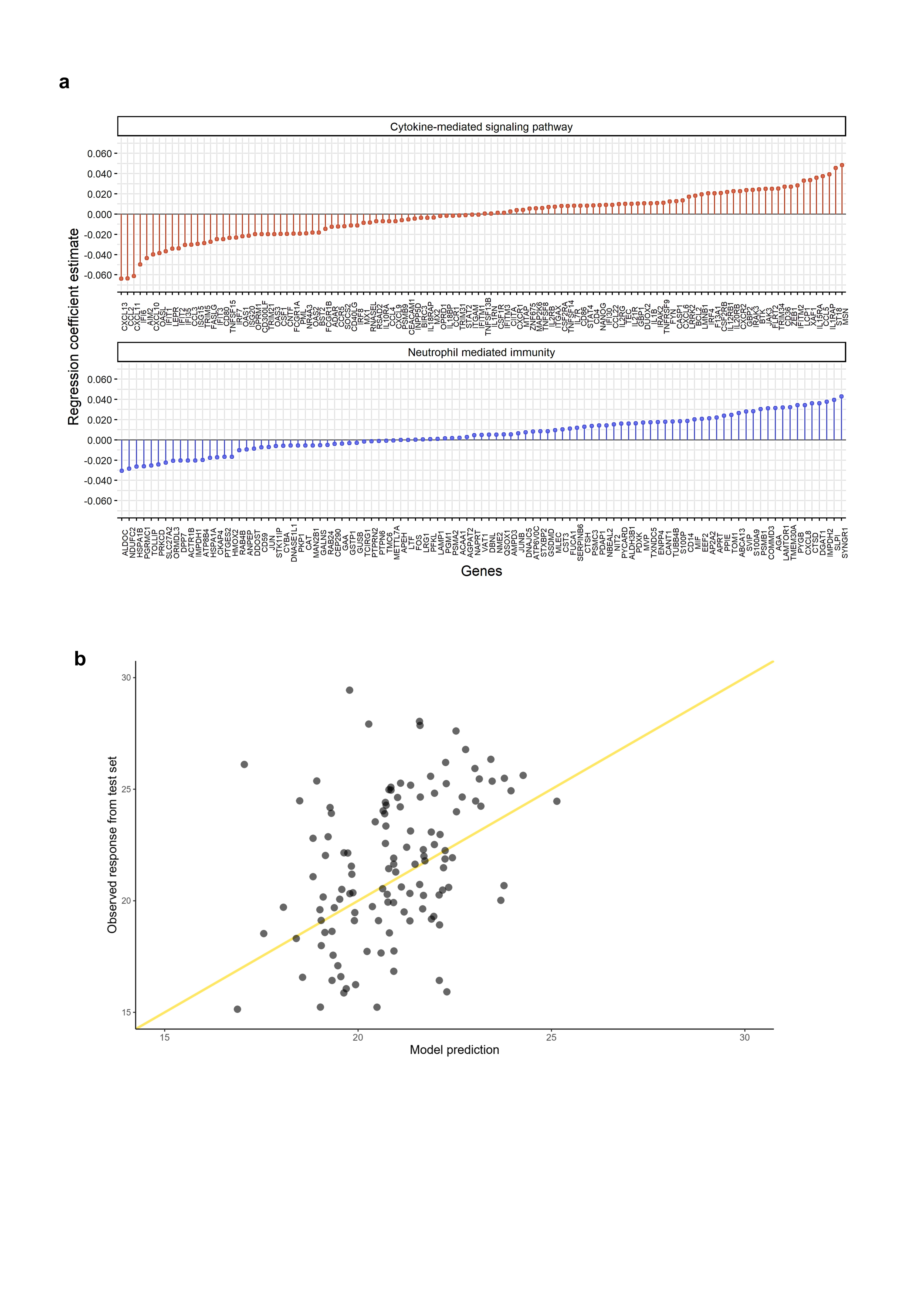

### ExtendedDataFig.3.tiff

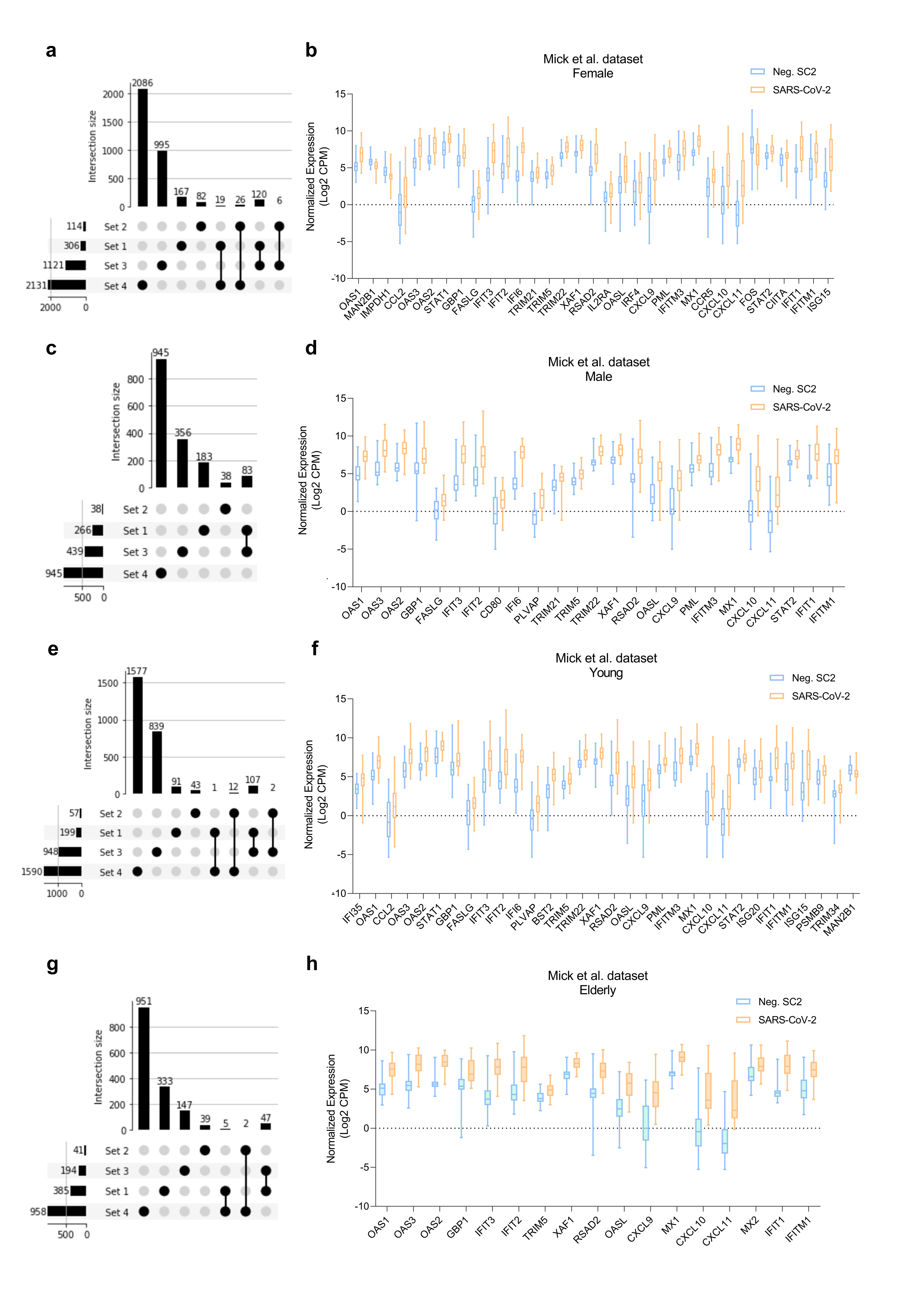

### ExtendedDataFig.4.tiff

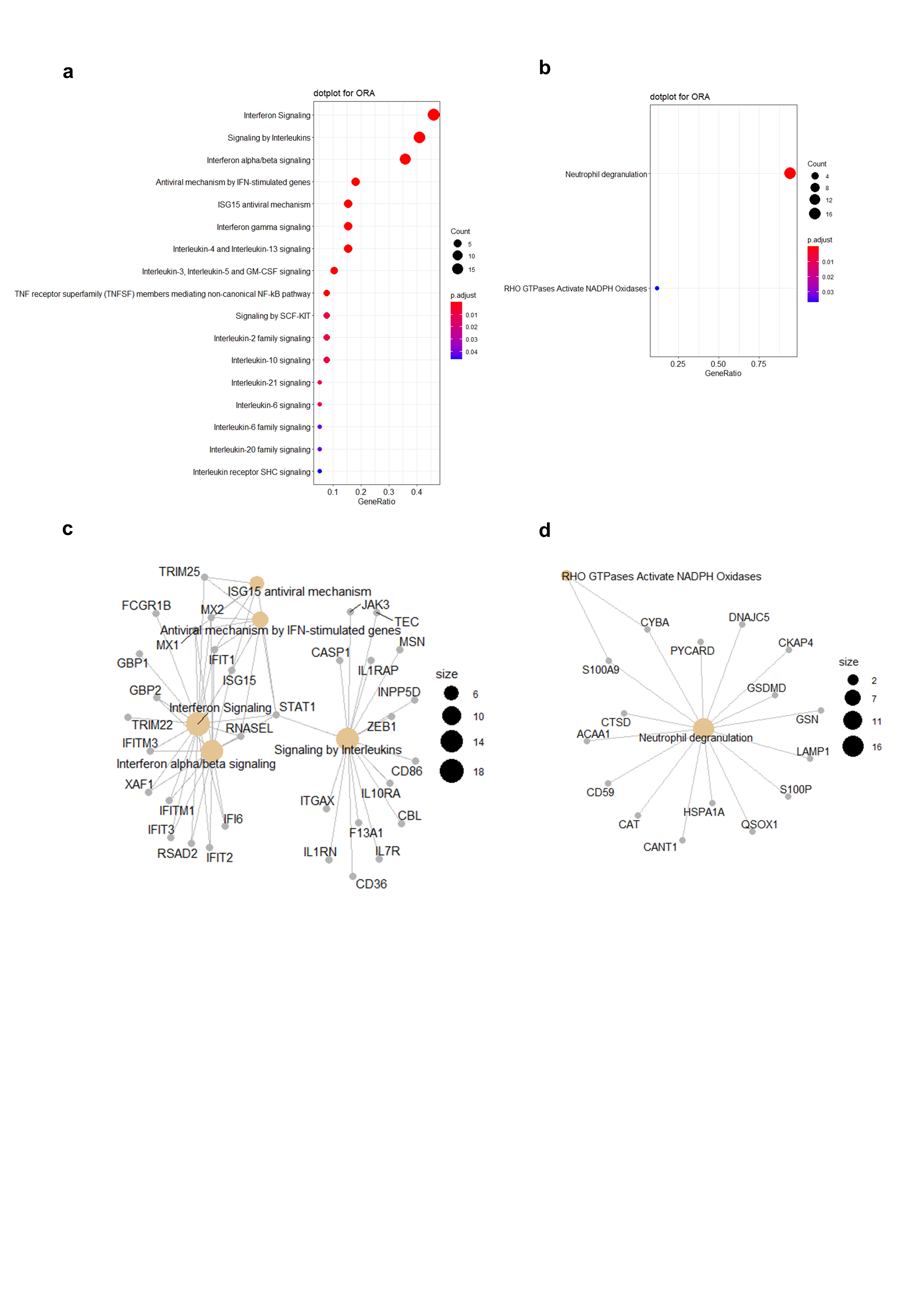

### ExtendedDataFig.5.tiff

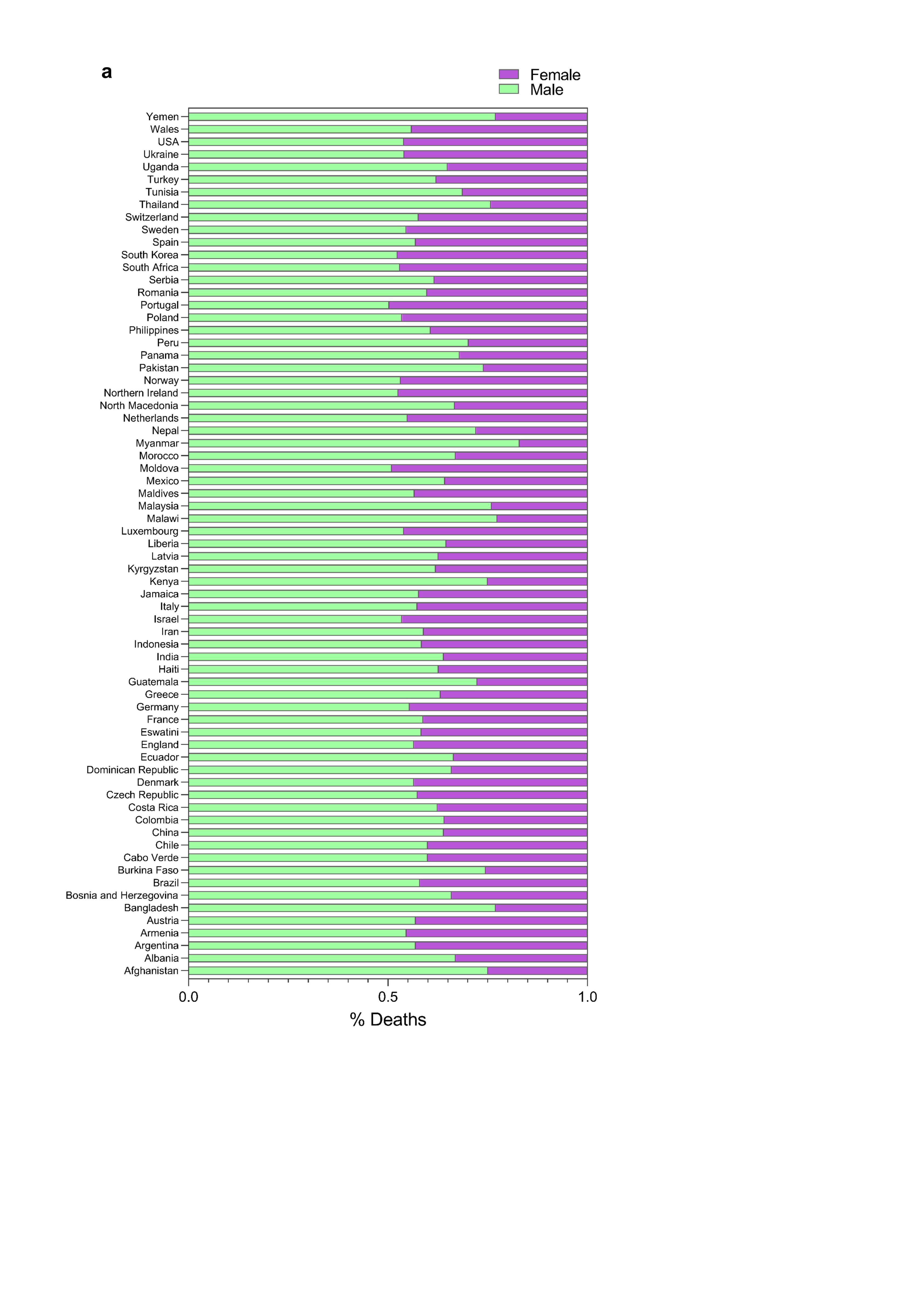

### ExtendedDataFig.6.tiff

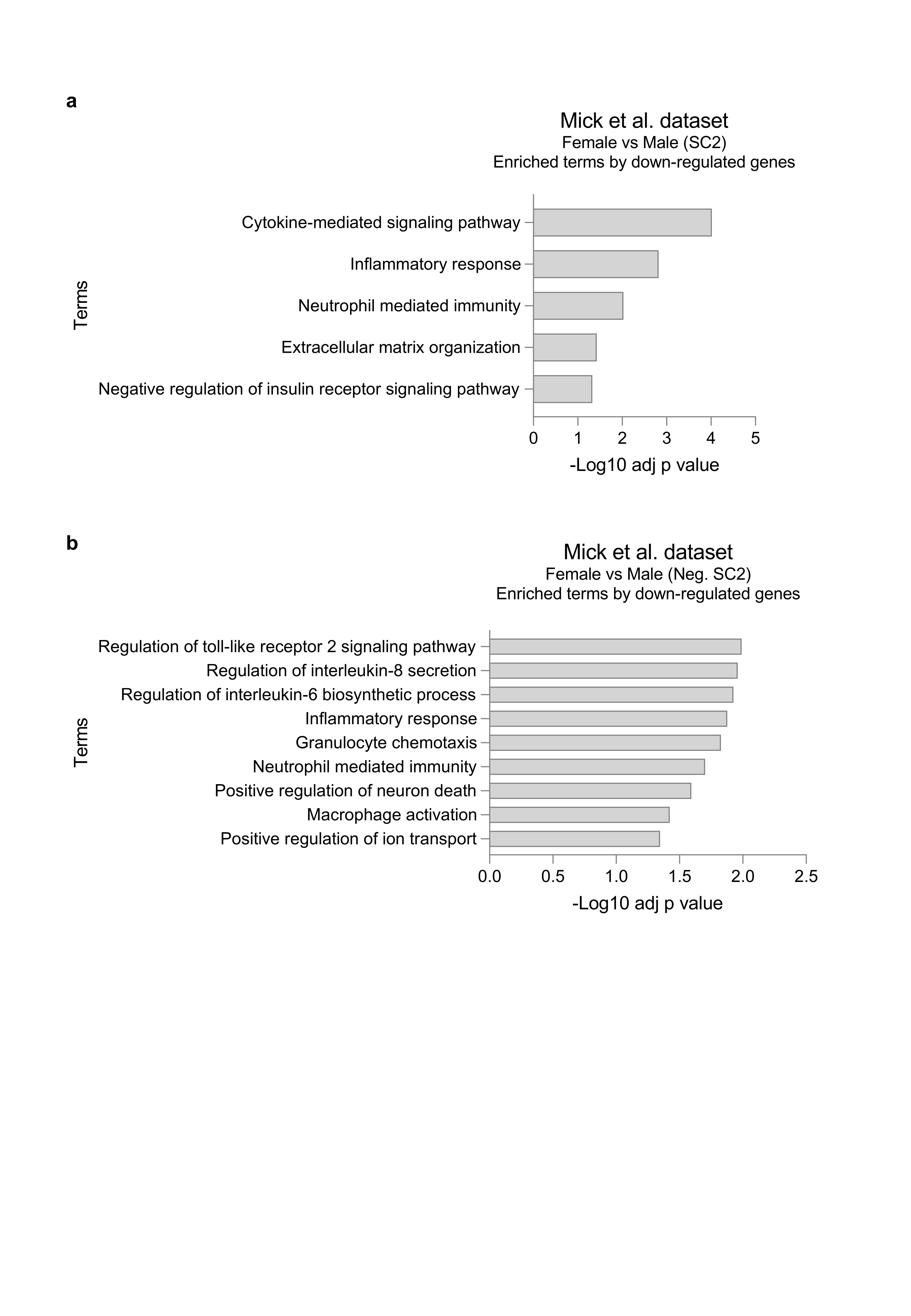

### ExtendedDataFig.7.tiff

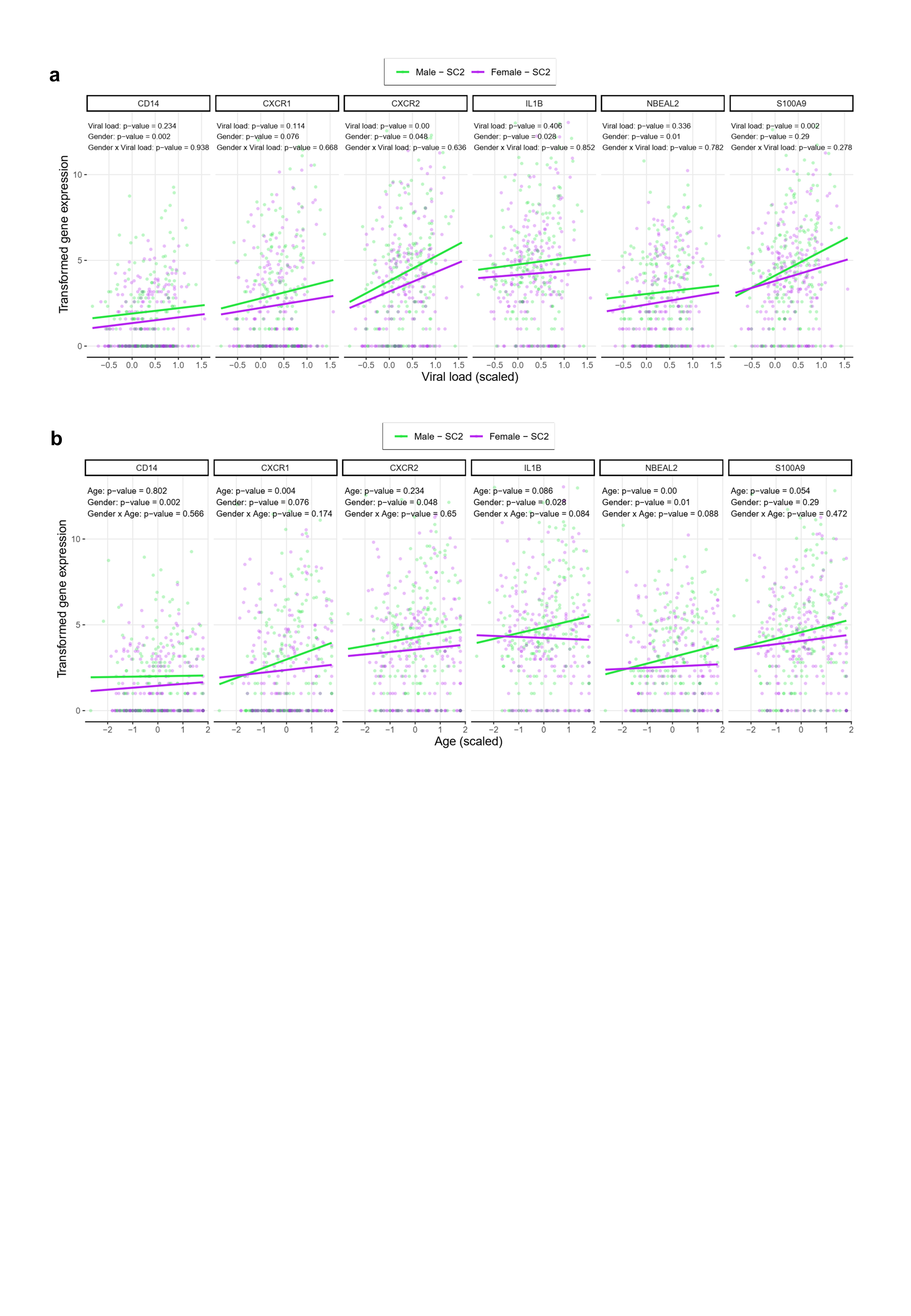

### ExtendedDataFig.8.tiff

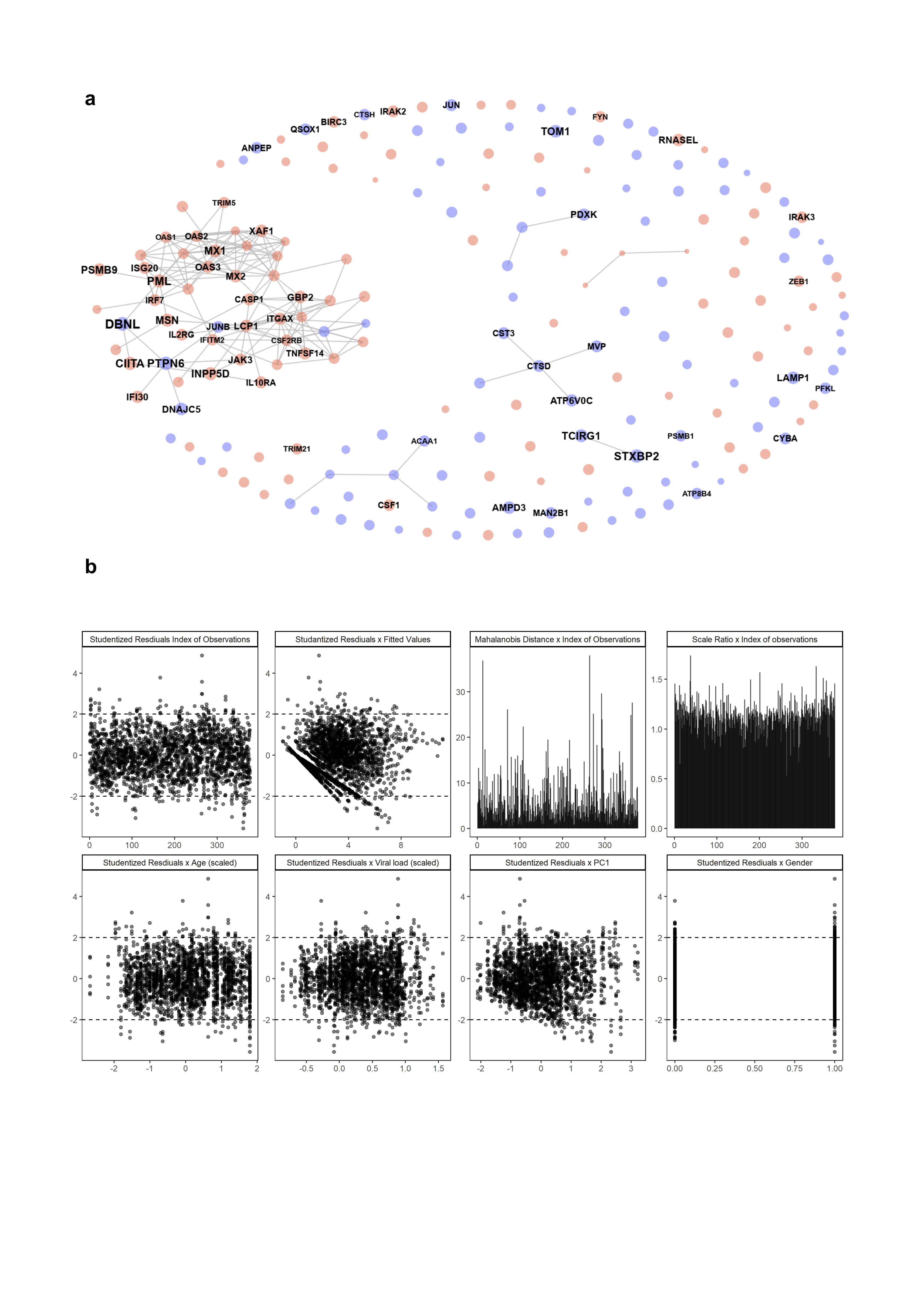
